# Psychometric Properties and Continuous National Norms of the Parent-Report Concise Health Risk Tracking Assessment: Charting Pediatric Suicide Risk Across Age and Sex

**DOI:** 10.64898/2026.09.21.26363585

**Authors:** Halle Deericks, Eric A. Youngstrom, Mindy Westlund Schreiner

**Affiliations:** Center for Suicide Prevention & Research, The Abigail Wexner Research Institute at Nationwide Children’s Hospital, Columbus, OH; Institute for Mental & Behavioral Health Research, The Abigail Wexner Research Institute at Nationwide Children’s Hospital, Columbus, OH; Department of Psychiatry & Behavioral Health, The Ohio State University, Columbus, OH; Behavioral Health, Nationwide Children’s Hospital, Columbus, OH

**Author notes:** Corresponding Author: Mindy Westlund Schreiner, 444 Butterfly Gardens Drive, Columbus, OH 43215.

**Keywords:** Concise Health Risk Tracking (CHRT), suicide risk, parent-report, normative data

## Abstract

**Objective:** The Concise Health Risk Tracking (CHRT) assessment has utility in suicide risk prediction in adults and adolescents. Because multi-informant assessment is the gold standard in pediatric populations, a parent report version of the CHRT may enhance evaluation of youth suicide risk. We adapted the self-report version of the 16-item CHRT as a parent report version (CHRT-PR_16_) and evaluated the psychometric properties and continuous age- and sex-specific norms in a nationally representative sample of parents.

**Method:** Parents of youth aged 6-18 years (N=2,236; 2,235 in sex-stratified analyses) completed the CHRT-PR_16_, enabling evaluation of both the 16- and 14-item (CHRT-PR_14_) version. Psychometric analyses included internal consistency, factor structure, Item Response Theory, measurement invariance, criterion validity, and continuous norming using Generalized Additive Models for Location, Scale and Shape (GAMLSS).

**Results:** Internal consistency was excellent for both CHRT-PR_16_ and CHRT-PR_14_ (both *ω_t_* =.98, *α*=.96), although general factor variance was higher for CHRT-PR_14_ (*ω_H_*=.83) than CHRT-PR_16_ (*ω_H_*=.68). Mokken scale analysis confirmed zero monotonicity violations for CHRT-PR_14_. CHRT-PR_16_ showed a clinically trivial edge in criterion correlations with other measures of psychopathology. In GAMLSS modeling, a zero-inflated negative binomial distribution achieved optimal fit for both scales. Score trajectories diverged by age and sex around age 10, with males scoring slightly higher at upper percentiles.

**Conclusions:** CHRT-PR_16_ and CHRT-PR_14_ demonstrate strong psychometric support, essential unidimensionality, measurement invariance, and optimal GAMLSS fit. Age- and sex-specific percentile lookup tables provide population-level benchmarks to guide measurement-based care, symptom tracking, and suicide risk stratification in pediatric settings.

---

Over the past two decades, suicide rates among children and adolescents in the United States have increased substantially (Centers for Disease Control and Prevention, 2024). Across the United States, suicide is the second leading cause of death among children and adolescents aged 6–18 years. Furthermore, the number of suicides among children aged 6–12 years increased by approximately 83.6%, rising from 73 deaths in 2001 to 134 deaths in 2024. Due to this public health crisis, it is important that we have valid and reliable suicide-related measures that facilitate early detection of suicidal ideation and behavior in youth. Brief self-report measures are particularly valuable because they reduce respondent burden and support the timely identification of those who may be at risk (Batterham et al., 2015).

One of the most widely used suicide measures is the clinician-administered Columbia– Suicide Severity Rating Scale (C-SSRS), which was developed to distinguish between suicidal ideation and suicidal behavior (Posner et al., 2011). However, administration of the C-SSRS requires trained clinicians and dedicated clinical time, resources that may not be available in healthcare or research settings where time is limited (Mayes et al., 2023). The Concise Health Risk Tracking Self-Report (CHRT-SR) was developed as a self-report measure that could be repeated to detect changes in suicide risk over time and to facilitate assessment in both clinical and research settings (Trivedi et al., 2011). The CHRT-SR assesses multiple dimensions associated with suicide risk, including hopelessness, self-worth, pessimism about the future, perceived social support, and suicidal thoughts and plans. In a study of adolescents, Mayes et al. (2023) compared the CHRT-SR with the C-SSRS and found that both measures were effective in assessing and detecting suicide risk. However, the CHRT-SR provided additional information regarding suicide risk factors related to an individual’s propensity for suicidal behavior, suggesting that it may complement clinician-administered assessments by capturing broader psychological risk factors.

Multiple versions of the CHRT-SR have been validated, including the 7-item (CHRT-SR_7_), 12-item (CHRT-SR_12_), and 14-item and (CHRT-SR_14_) versions (Ostacher et al., 2015; Mayes et al., 2020). More recently, the 16-item version (CHRT-SR_16_) has also been validated (Trombello et al., 2023). The 16-item version consists of an additional two items that assess irritability. Despite the multiple versions of this measure, they have so far been limited to self-report versions. Multiple informants can be particularly helpful in assessing youth as parent and youth credibility were associated with different characteristics, with little overlap in their predictors, supporting the use of multiple informants in clinical assessment (Youngstrom et al., 2011). According to the *situational specificity* hypothesis, as described by Achenbach and colleagues, each informant provides accurate yet distinct information about a youth’s functioning based on the unique situations and contexts in which they observe the youth (Achenbach et al., 1995; De Los Reyes et al., 2023). Consistent with this hypothesis, incorporating parent report alongside youth self-report may provide a more comprehensive assessment of clinically relevant behavior and symptoms.

There also are clinical scenarios where parent report may be a first or preferred source of information. Parents can provide reliable and valid information about youth behavior and mood when children are often too young to have sufficiently developed meta-cognition or insight to have perspective on how their behavior affects others. Parents also are more likely to read sufficiently well to complete scales and checklists about younger children who themselves might not have the requisite skills. For these reasons all major behavioral rating scales have developed and validated parent report measures of youth behavior to extend below the age ranges for which corresponding youth self-report scales have been validated (e.g., Achenbach & Rescorla, 2001; Gadow, 2016; Sprafkin et al., 2002). In addition, it is usually the parent who initiates referrals for outpatient mental health services. Parents also may sometimes seek services for their youth even when the youth does not perceive the need for services or is actively avoiding help. In short, there are a variety of clinical needs that a parent-report measure of risk of youth self-injury could address.

To extend the assessment of suicide risk across informants, our study team adapted the CHRT-SR_16_ into a parent-report measure (CHRT-PR_16_). We then established normative data by administering this assessment to a nationally representative sample as part of a larger public domain measurement-based care norming project — BENCHMARK 2025 (osf.io/yxf8z). Normative benchmarks describe whether a given score reflects typical or atypical functioning relative to the general population and could inform clinical decision making as it relates to symptom severity or impairment (Achenbach, 2001). In the context of child mental health and general functioning, national norms are especially valuable for tracking individual progress against population-level, non-clinical reference points, informing treatment plans, and flagging meaningful deterioration or improvement. Specifically, we examined the psychometrics and normative data for both the 16- and 14-item versions of the CHRT-PR, as the 14-item version has a stronger youth evidence base for prospective risk stratification for suicidal behavior (Mayes et al., 2020).

## Method

### Procedure and Participants

This study was approved by the Institutional Review Board at Nationwide Children’s Hospital. The survey agency YouGov administered selected measures to parents. YouGov has a global panel of over 29 million registered members, many of whom are located in the U.S., enabling the company to conduct nationally representative surveys. YouGov collected and cleaned all responses and applied stratification based on key demographics (child sex, race, ethnicity, and parent education level) to align with the 2023 American Community Survey (ACS) one-year estimates, resulting in a sample representative of the U.S. population on key demographics.

To evaluate test-retest dependability and establish clinical significance benchmarks, a pre-registered subsample of parents completed a repeat administration of the survey battery approximately two weeks following baseline assessment.

### Measures

#### Concise Health Risk Tracking Scale Parent-Report (CHRT-PR) 16- and 14-Item Versions

Adapted from the youth self-report (CHRT-SR_16_; Trivedi et al., 2011), the CHRT-PR _16_ is a 16-item scale that assesses parent-reported suicide severity and risk in their child. Each item is rated on a five-point Likert scale (0 = *strongly disagree* to 4 = *strongly agree*), with total scores ranging from 0 to 56. The CHRT-PR_16_ and CHRT-PR_14_ differ in that the 16-item version includes two questions assessing irritability. Of note, these items were inserted into the 14-item set rather than being appended at the end, appearing as items #12 and #13 in the sequence.

#### Criterion Validity

In addition to the CHRT-PR _16_, participants in the normative sample also provided demographic information and completed other parent-report short scales frequently used in measurement-based care. We selected a subset likely to show convergent validity with the CHRT-PR _16_ scores, including the Very Quick Inventory of Depressive Symptomatology (5-item version; (VQIDS5; De La Garza et al., 2017), the Pediatric Symptoms Checklist-17 (PSC-17; Gardner et al., 1999), the Nationwide Quality of Life Scale (NQLS; Covarrubias & Fristad, 2025)), and the N-Gauge, a parent-rated global impairment scale (McClellan et al., 2025). The PSC-17 provides a total score and three factor-informed subscales: Internalizing, Externalizing, and Attention Problems. We anticipated that CHRT scores would show the highest correlation with the VQIDS5 and the Internalizing subscale of the PSC-17.

## Data Analysis

### Factor Structure

Prior to conducting the EFA, the suitability of the data for factor analysis was assessed using the Kaiser-Meyer-Olkin (KMO) measure of sampling adequacy and Bartlett’s test of sphericity. Exploratory factor analyses (EFA) were conducted to examine the underlying factor structure of the CHRT-PR and evaluate the dimensionality of its items using the *lavaan* package (Rosseel, 2012). The number of factors was determined based on examination of eigenvalues, the scree plot, optimal coordinates, and parallel analysis, per preregistration. Initial analyses were conducted using principal axis factoring based on a polychoric correlation matrix. One-, two-, and three-factor solutions were examined to evaluate alternative dimensional structures.

A bifactor confirmatory factor analysis (CFA) and a second-order (hierarchical) factor model was also conducted to evaluate the presence of a general factor and specific factors underlying the CHRT-PR items. Because the items were ordinal, the bifactor CFA was conducted using the weighted least squares mean- and variance-adjusted (WLSMV) estimator, with CHRT-PR_16_ and CHRT-PR_14_ items specified as ordered categorical variables. Model fit was evaluated using the scaled *X^2^*, RMSEA, CFI, TLI, and SRMR. Standardized factor loadings were examined to evaluate the strength of the relationships between individual items and the general and specific factors. The final factor structure was determined based on model fit, factor loadings, and theoretical interpretability.

### Internal Consistency and Measurement Precision

Internal consistency reliability of the CHRT-PR_16_ and CHRT-PR_14_ was assessed using Cronbach’s *α* (Cronbach, 1951) and McDonald’s *ω* (McDonald, 1999). Dimensionality was evaluated using a Schmid-Leiman omega analysis and associated bifactor indices to assess the strength of the general factor and determine whether the measure demonstrated sufficient essential unidimensionality to support a unidimensional item response theory (IRT) model. Prior to IRT model estimation, monotonicity was evaluated using Mokken scale analysis.

A graded response model (GRM) was estimated to evaluate item functioning and measurement precision. The GRM was selected because all CHRT-PR_16_ and CHRT-PR_14_ items used a five-category ordinal response scale (0-4). Item discrimination and threshold parameters were examined, and item and test information functions were evaluated to assess measurement precision. Empirical reliability of the IRT-based scores was also estimated.

Prior to norm construction, item- and model-level measurement invariance were evaluated across child sex and age to confirm that CHRT-PR_16_ and CHRT-PR_14_ items function equivalently across these subgroups. Item-level differential item functioning (DIF) was assessed using iterative hybrid ordinal logistic regression (lordif; Choi et al., 2009), which combines an ordinal logistic regression DIF test with IRT-based trait estimation, iterating until no further items are flagged; an item was flagged when adding group membership and a group-by-trait interaction improved model fit by McFadden’s pseudo-*R²* ≥ .02. Age was evaluated across three developmentally-informed strata (6–9, 10–12, and 13–18 years) using pairwise comparisons.

Model-data fit of the GRM was further evaluated at the item- and person-level following Feuerstahler, Waller, and MacDonald’s (2020) applied framework: local item dependence via the Q3 statistic (Yen, 1984); flagged at |Q3| > .20), absolute item fit via the root-mean-square deviation between model-implied and observed item response functions (RMSD; Yamamoto et al., 2013; flagged at RMSD > .10), and person fit via the standardized Zh statistic (Drasgow et al., 1985; flagged at |Zh| > 3).

Model-level invariance was evaluated separately via a multi-group confirmatory factor analysis sequence, fit with the WLSMV estimator on ordered-categorical items, identical to the structural CFA procedure described above. For each facet (sex and age), three nested models were compared: a configural model (equivalent item-factor structure, no cross-group equality constraints), a metric model (equal item loadings across groups), and a scalar model (equal item loadings and thresholds across groups). Invariance at each step was evaluated using the change in comparative fit index (ΔCFI) between nested models, with ΔCFI ≤ .010 indicating that the added equality constraints did not meaningfully degrade model fit (Chen, 2007; Cheung & Rensvold, 2002) — the standard criterion for concluding a given level of invariance holds. Sex- and age-stratified analyses were run separately for the CHRT-PR_16_ and CHRT-PR_14_ item sets.

### Regression-Based Continuous Norming

We established age- and sex-referenced norms for the CHRT-PR_16_ and CHRT-PR_14_ using Generalized Additive Models for Location Scale and Shape (GAMLSS). Guided by continuous norming frameworks for highly skewed, zero-heavy clinical scales (Timmerman et al., 2021, 2026), we evaluated three discrete candidate distributions designed to handle count data with excess zeros: Negative Binomial Type II (NBI), Zero-Inflated Negative Binomial (ZINBI), and Zero-Adjusted Negative Binomial (ZANBI).

We used penalized B-spline (P-spline) smoothers on all candidate models to model non-linear, age-dependent changes in parameters. Specifically, the mean (µ) submodel was parameterized with a sex main effect, a penalized spline of age, and a sex-by-age interaction to allow developmental trajectories to vary dynamically between males and females. The dispersion (*σ*) submodel was also modeled using a penalized spline function of age to capture age-dependent changes in score variance, while we held constant the zero-inflation parameter (*ν* ), which represents the probability of structural zeros.

We selected our model using global and local fitness indices, including the Bayesian Information Criterion (BIC), worm plots to inspect normalized quantile residuals across age subgroups, and Q-statistics to detect localized misfit across age bands. The target calibration of the fitted models was verified against nominal empirical thresholds (5^th^, 10^th^, 25^th^, 50^th^, 75^th^, 90^th^, and 95^th^ percentiles). We also conducted an 80/20 train-test holdout validation evaluated separately by sex to ensure robust predictive validity.

### Clinical Significance and Reliable Change Benchmarks

Test-retest dependability was evaluated across the two-week retest subsample using absolute-agreement intraclass correlation coefficient, ICC(A,1), and Pearson correlations. These dependability estimates were subsequently used to calculate the 95% Reliable Change Index (RCI) following Jacobson and Truax’s framework (), establishing empirical thresholds for distinguishing true clinical improvement or deterioration from measurement error and short-term state variability. Additionally, the Minimally Important Difference (MID; half-*SD* heuristic) and Jacobson-Truax non-clinical recovery thresholds (*M + 2SD*) were derived to contextualize individual score changes. These benchmarks are shown in Table 4.

## Results

### Participant Characteristics

Parents of 2,252 children aged 6-18 completed at least some of the assessments with 2,236 completing all items of the CHRT-PR_16_. For analyses that examined incorporated sex differences, 2,235 participants were included due to one child being identified as intersex. This included differential item functioning, measurement invariance, and GAMLSS analyses. Based on parent report, children were distributed across the 6-18 age range, with 47.5% of children being female, 75.3% White, 8.8% Black or African American, 3.7% Asian, and 16.4% Hispanic, Latino, or Spanish Origin. Nearly all parents (97.6%) had obtained a high school diploma, and 47.5% had obtained a bachelor’s degree or higher. Median household income was between $70,000 and $79,999 annually. Key demographics are summarized in Table 1. For the repeat administration of assessments, 170 parents completed the survey battery a second time two weeks following baseline.

**Table 1.**
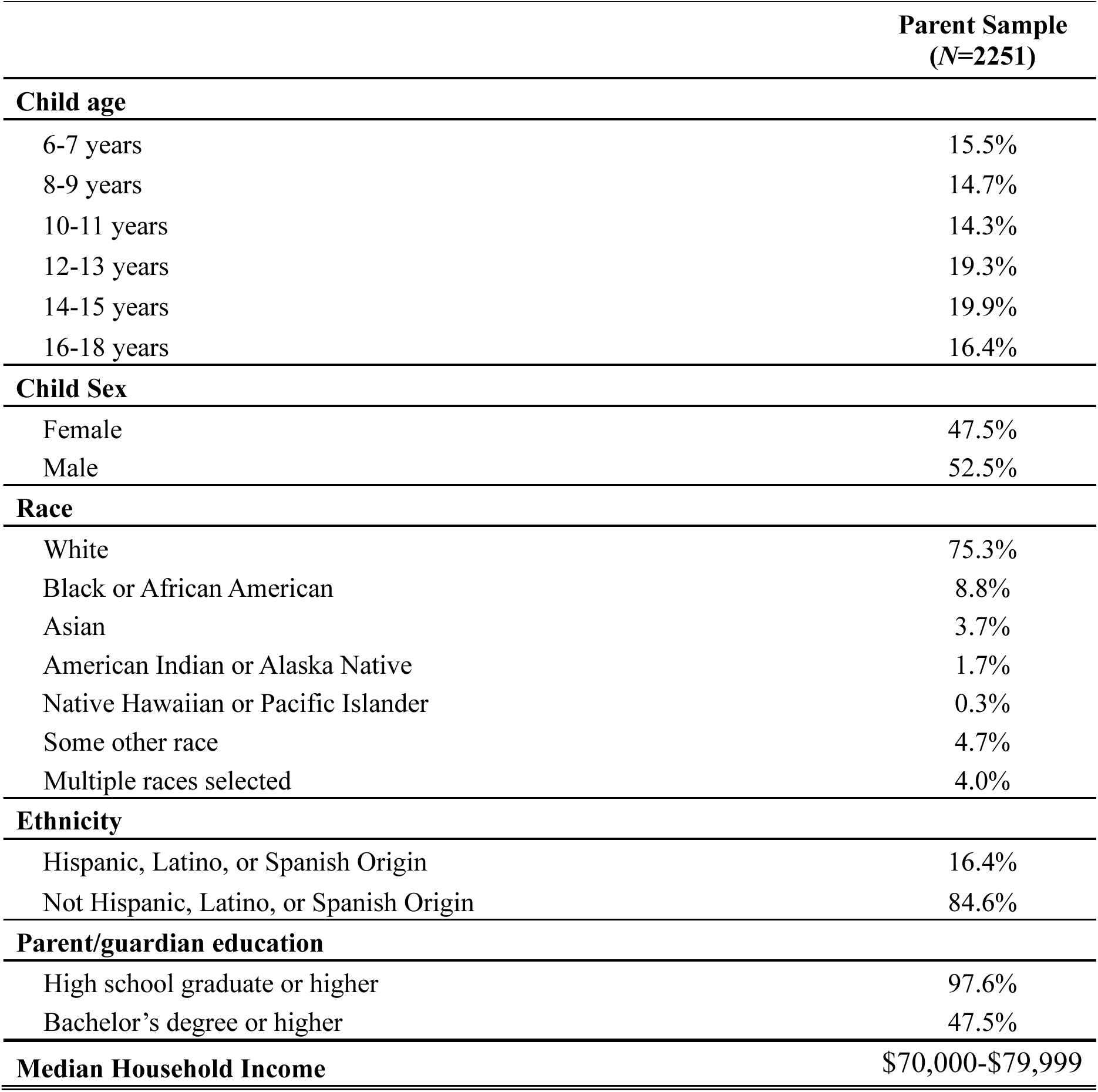
Participant demographic characteristics.

|  | Parent Sample<br>(N=2251) |
| --- | --- |
| **Child age** |  |
| 6-7 years | 15.5% |
| 8-9 years | 14.7% |
| 10-11 years | 14.3% |
| 12-13 years | 19.3% |
| 14-15 years | 19.9% |
| 16-18 years | 16.4% |
| **Child Sex** |  |
| Female | 47.5% |
| Male | 52.5% |
| **Race** |  |
| White | 75.3% |
| Black or African American | 8.8% |
| Asian | 3.7% |
| American Indian or Alaska Native | 1.7% |
| Native Hawaiian or Pacific Islander | 0.3% |
| Some other race | 4.7% |
| Multiple races selected | 4.0% |
| **Ethnicity** |  |
| Hispanic, Latino, or Spanish Origin | 16.4% |
| Not Hispanic, Latino, or Spanish Origin | 84.6% |
| **Parent/guardian education** |  |
| High school graduate or higher | 97.6% |
| Bachelor's degree or higher | 47.5% |
| **Median Household Income** | \$70,000-\$79,999 |

### Factor Structure

#### CHRT-PR_16_

The KMO measure of sampling adequacy indicated that the data were suitable for factor analysis, KMO = .94, exceeding the recommended minimum of .60. Bartlett’s test of sphericity was significant, *χ2*(120) = 43,273.5 *p* < .0005, indicating that the item correlations were sufficiently large to support factor analysis.

To evaluate the underlying factor structure of the CHRT-PR_16_, one-, two-, and three-factor EFA solutions were examined. The one-factor solution demonstrated poor fit, with a Tucker-Lewis Index (TLI) of .677 and an RMSEA of .228, 90% CI [.224, .231]. The BIC was 11,374.16. A two-factor solution demonstrated improved fit, with a TLI of .798 and an RMSEA of .180, 90% Cl [.176, .184]. The BIC was 5,851.59. The three-factor solution demonstrated further improvement, with a TLI of .852 and an RMSEA of .154, 90% CI [.150, .158]. The BIC decreased further to 3,480.29, indicating better relative fit compared to the one- and two-factor solutions. The second-order model was not empirically identifiable.

Given the relatively poor absolute fit of the EFA solutions, a bifactor CFA was conducted to further evaluate the dimensionality of the CHRT-PR_16_. The bifactor model demonstrated good fit to the data, *χ2(88)* = 822.75 p < .001, CFI = .997, TLI = .996, RMSEA = .061, 90% CI [.057, .065], and SRMR = .034. Schmid-Leiman EFA decomposition was used to calculate omega hierarchical (*ω*H*) =* .68, suggesting that 68% of the reliable variance in total scores was attributable to the general factor.

#### CHRT-PR_14_

The same analytic approach was used to evaluate the factor structure of the CHRT-PR_14_. The KMO measure of sampling adequacy indicated that the data were suitable for factor analysis, KMO = 0.94. Bartlett’s test of sphericity was significant, *χ2*(91) = 37,932.7 *p* < .0005, indicating that the item correlations were sufficiently large to support factor analysis.

One-, two-, and three-factor EFA solutions were examined. The one-factor solution demonstrated poor fit, with a Tucker-Lewis Index (TLI) of .733 and an RMSEA of .223, 90% CI [.219, .227]. The BIC was 8,035.24. A two-factor solution demonstrated improved fit, with a TLI of .812 and an RMSEA of .187, 90% Cl [.182, .191]. The BIC was 4,562.30. The three-factor solution demonstrated further improvement, with a TLI of .899 and an RMSEA of .137, 90% CI [.132, .142]. The BIC was 1,841.35.

Given the relatively poor fit of the EFA solutions, a bifactor CFA was conducted to further evaluate the dimensionality of the CHRT-PR_14_. However, the model failed to achieve convergence due to empirical underidentification, and its fit statistics were therefore not interpreted. Schmid-Leiman EFA decomposition was used to calculate omega hierarchical (*ω*_H_*) =* .83, suggesting that 83% of the reliable variance in total scores was attributable to the general factor.

### Internal Consistency and Measurement Precision

#### CHRT-PR_16_

The CHRT-PR_16_ demonstrated good internal consistency (Cronbach’s *α*= .96; McDonald’s *ω*= .98). Given that Schmid-Leiman analysis indicated a strong general factor, with omega hierarchical (*ω*H*) =* .68, these findings provided support for the use of a unidimensional IRT model. Mokken scale analysis indicated that the items demonstrated adequate to strong scalability, with item-level scalability coefficients ranging from *H*_i_ = .48 to .58. Only a minor violation of monotonicity was identified between items 10 and 12 (*v*_i_/AC = .01). Given the negligible magnitude of this violation, the items were considered to satisfy the monotonicity assumption for GRM estimation.

In the GRM estimation, item discrimination parameters ranged from *a* = 1.25 to 6.04, indicating moderate to very high discrimination across items (see Supplemental Table S1). Absolute item-level model fit was excellent across all 16 items, with RMSD values ranging from .007 to .058 (see Supplemental Table S2), confirming zero items exceeded the established .10 misfit threshold (Feuerstahler et al., 2020). Threshold parameters (*b*1 – *b*4) ranged from –0.76 to 2.99, with thresholds generally increasing across response categories, indicating appropriate ordering of the response options. Evaluation of local item dependence revealed that 20.8% of item pairs exceeded **|***Q*_3_**| >** .20 (see Supplemental Table S3), with the highest residual dependence occurring between the two irritability items (#12 and #13; *Q_3_* = .660; see Supplemental Table S4). Conditional reliability was ≥ .80 across a broad range of the latent trait (θ = -0.76 to 3.68) with the highest reliability approaching .98 in the middle-to upper range of the trait. Empirical reliability of the IRT-based scores was 0.88, indicating high reliability of the estimated trait scores. The simple unit-weighted total score correlated very highly with the IRT-based EAP score for the CHRT-PR_16_ (*r* = .906).

#### CHRT-PR_14_

The same analytics approach was used to evaluate the CHRT-PR_14_. This measure demonstrated good internal consistency (Cronbach’s *α*= .96; McDonald’s *ω*= .98). Similar to the CHRT-PR_16_, Schmid-Leiman analysis indicated a strong general factor, with omega hierarchical (*ω*_H_) *=* .83, providing support for the use of a unidimensional IRT model. Mokken scale analysis indicated that the items demonstrated adequate to strong scalability, with item-level scalability coefficients ranging from *H*_i_ = .45 to .62. Given no violation of monotonicity, the items were considered to satisfy the assumption for GRM estimation.

In the GRM estimation, item discrimination parameters ranged from *a* = 1.07 to 6.22, indicating moderate to very high discrimination across items (see Supplemental Table S5). Similarly, absolute item fit for the CHRT-PR_14_ GRM was strong across all 14 items, with RMSD values ranging from .006 to .086 (see Supplemental Table S6). Threshold parameters (*b*1 – *b*4) ranged from –0.84 to 3.36, with thresholds generally increasing across response categories, indicating appropriate ordering of the response options. For the CHRT-PR_14_, 20.9% of item pairs exceeded *Q*_3_**| >** .20 (see Supplemental Table S3). Remaining local dependence was concentrated within distinct clinical doublets, such as impulsivity (items #10 and 11, *Q_3_* = .635; see Supplemental Table S7). Notably, eliminating items 12 and 13 removed the prominent irritability secondary factor, which had previously absorbed general trait variance in the 16-item version. This removed a major source of multidimensional noise, allowing the general suicide risk factor to account for a markedly higher proportion of reliable variance (*ω*_H_ = .83 versus .68). Conditional reliability was ≥ .80 across a broad range of the latent trait (θ = -0.54 to 3.65) with the highest reliability approaching .98 in the middle-to upper range of the trait. Empirical reliability of the IRT-based scores was 0.85, indicating high reliability of the estimated trait scores. Similar to the CHRT-PR_16_, the simple unit-weighted total score correlated very highly with the IRT-based EAP score (CHRT-PR_14_: *r* = .905), indicating that the more clinically interpretable raw sum provides an essentially equivalent approximation of the more computationally intensive IRT-derived trait estimate for either scale length. Thus we carried the unit-weighted score forward into the development of norms and the evaluation of criterion validity.

### Differential Item Functioning and Measurement Invariance

To evaluate whether CHRT-PR_16_ and CHRT-PR_14_ items function equivalently across child sex and age, a prerequisite for interpreting a single total score the same way across subgroups, item- and model-level invariance were tested for both facets. Item-level DIF was evaluated using iterative hybrid ordinal logistic regression (lordif; McFadden ΔR² ≥ .02 flag threshold), and confirmatory factor models were fit at the configural, metric (equal loadings), and scalar (equal thresholds) levels, with ΔCFI ≤ .010 as the invariance criterion. Age was tested across three strata: 6–9 (middle childhood), 10–12 (pubertal transition), and 13–18 (adolescence).

No items were flagged for DIF on either facet, for either scale version. Model-level invariance was equally clean: for sex, both scales’ configural models fit well (CHRT-PR_16_: CFI = .977; CHRT-PR_14_: CFI = .983) with negligible degradation at the metric (ΔCFI = −.001 and 0.000, respectively) and scalar (ΔCFI = +.001 and 0.000) levels. Age showed the identical pattern (configural CFI = .977 and .984; metric ΔCFI = −.001 for both; scalar ΔCFI = +.001 for both). All four invariance sequences (2 scales × 2 facets) held through the strictest (scalar) level, corroborated by the absence of any item-level DIF flag.

## Criterion Validity

CHRT-PR_16_ correlated more strongly than CHRT-PR_14_ with every external criterion examined: depression severity, functional impairment, and mental health service utilization alike (*r* = .17 to .67 across measures); see Figure 1. This pattern reached statistical significance for most criteria given the large sample, but the magnitude of every difference was negligible (Cohen’s *q* < .08; see Table 2). The two versions are therefore practically equivalent in criterion-related validity despite the consistent numerical edge of the CHRT-PR_16_. Furthermore, cross-validated scoring-method sensitivity analyses on the 2-week retest holdout (N = 170) confirmed that simple unit-weighted raw scores performed equivalently to IRT EAP theta scores across all 12 external criteria, with no statistically or clinically meaningful advantage for IRT scoring (see Supplemental Table S8).

**Figure 1.**
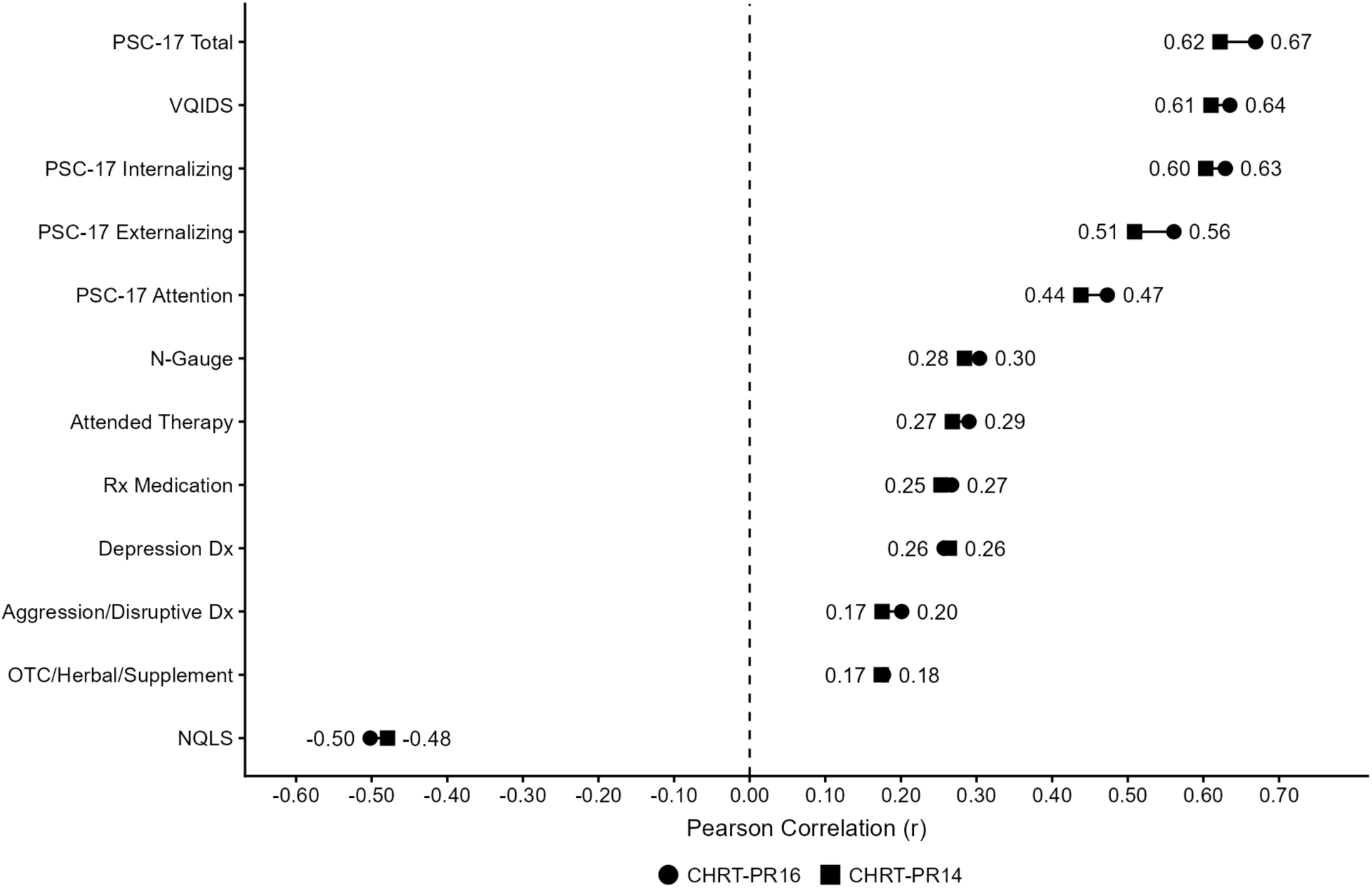
Concurrent Criterion-Related Validity Comparisons Between the CHRT-PR16 and CHRT-PR14 Across External Clinical Measures. **Note.** Pearson correlation coefficients (*r*) comparing unit-weighted total scores for the 16-item (CHRT-PR16) and 14-item (CHRT-PR14) parent-report scales against 12 concurrent external clinical criteria in the complete baseline sample (*N* = 2,227–2,239). CHRT-PR_16_ and CHRT-PR_14_ total scores correlated *r* = .983 with each other. Pairwise statistical comparisons between dependent overlapping correlation coefficients were conducted using Steiger’s (1980) *z*-test, with *p*-values adjusted for multiple comparisons using the Holm–Bonferroni procedure. Effect sizes for differences between correlation coefficients are reported as Cohen’s *q*, where *q* = .10, .30, and .50 denote small, medium, and large effect sizes respectively. While the CHRT-PR16 exhibited statistically significantly higher correlations across most criteria due to the large sample size, all difference effect sizes were clinically trivial (*q* < .08). Absolute correlations *r* ≥ .07 are statistically significant at *p* < .001 (two-tailed). *Abbreviations:* PSC-17 = Pediatric Symptom Checklist-17; VQIDS = Very Quick Inventory of Depressive Symptomatology (5-item parent proxy); NQLS = Nationwide Quality of Life Scale (reverse-scored); N-Gauge = Parent-Rated Global Impairment Scale; Rx = Prescription; Dx = Diagnosis; OTC = Over-the-Counter/Herbal/Supplement.

**Table 2.** Concurrent Criterion-Related Validity Comparisons Between CHRT-PR_16_ and CHRT-PR_14_ Across External Clinical Measures.

| Criterion Measure | *r*<sub>PR16</sub> | *r*<sub>PR14</sub> | Steiger's *z* | *p* (Holm) | Cohen's *q* |
| --- | --- | --- | --- | --- | --- |
| PSC-17 Total | .669 | .622 | 15.83 | < .001 | .080 |
| PSC-17 Externalizing | .561 | .509 | 15.98 | < .001 | .073 |
| PSC-17 Attention | .473 | .438 | 10.13 | < .001 | .044 |
| PSC-17 Internalizing | .629 | .603 | 8.40 | < .001 | .041 |
| VQIDS (Depression Severity) | .635 | .610 | 8.18 | < .001 | .040 |
| Attended Therapy | .290 | .268 | 5.93 | < .001 | .024 |
| Aggression / Disruptive Dx | .201 | .175 | 6.64 | < .001 | .026 |
| NQLS (Quality of Life) | -.502 | -.479 | -6.83 | < .001 | .030 |
| N-Gauge (Global Impairment) | .304 | .284 | 5.24 | < .001 | .021 |
| Rx Medication | .267 | .252 | 3.90 | < .001 | .016 |
| Depression Dx | .257 | .264 | -1.72 | .173 | .007 |
| OTC / Herbal / Supplement | .177 | .174 | 0.77 | .445 | .003 |

*Note.* Pearson correlation coefficients (*r*) comparing unit-weighted total scores for CHRT-PR<sub>16</sub> and CHRT-PR<sub>14</sub> against 12 concurrent external clinical criteria in the complete baseline sample (*N* = 2,227–2,239). CHRT-PR<sub>16</sub> and CHRT-PR<sub>14</sub> correlate *r* = .983 with each other. Pairwise statistical comparisons between dependent overlapping correlation coefficients were conducted using Steiger's (1980) *z*-test, with *p*-values adjusted for multiple comparisons using the Holm–Bonferroni procedure. Cohen's *q* values of .10, .30, and .50 denote small, medium, and large effect sizes, respectively. Absolute correlations *r* ≥ .07 are statistically significant at *p* < .001 (two-tailed).
*Abbreviations:* PSC-17 = Pediatric Symptom Checklist-17; VQIDS = Very Quick Inventory of Depressive Symptomatology; NQLS = Nationwide Quality of Life Scale (reverse-scored); N-Gauge = Parent-Rated Global Impairment Scale; Rx = Prescription; Dx = Diagnosis; OTC = Over-the-Counter.

### Regression-Based Continuous Norming

#### Model Fit and Selection

The three-parameter ZINBI distribution was the optimal model for both the 16- and 14-item versions of the CHRT-PR. ZINBI produced the lowest (most optimal) BIC values for both the 14-item (*BIC* = 13,938.94) and 16-item (*BIC* = 14,992.40) item versions. Under both scales, the NBI model failed on residual skewness (*p* ≤ .01), reflecting a severe misfit due to unmodeled zero-inflation. While ZANBI showed comparable diagnostic performance to ZINBI on both scales, it was outperformed by ZINBI on BIC (see Supplemental Table S9 and Supplemental Figures S1 and S2).

### GAMLSS Residual Diagnostics and Scale Justification

Both the CHRT-PR_16_ and CHRT-PR_14_ ZINBI models resolved residual skewness cleanly (CHRT-PR_16_ *Z_3_ p =* .61; CHRT-PR_14_ *Z_3_ p* = .77) but retained a significant residual kurtosis misfit (CHRT-PR_16_ *Z_4_ p <* .001; CHRT-PR_14_ *Z_4_ p* = .01). For the CHRT-PR_16_ model, this misfit was most concentrated in the 8-, 9-, 15-, and 17-year-old age bands. For the CHRT-PR_14_ model, it was most concentrated in the 9-year-old age band, with secondary flags at the 15- and 17-year-old age bands. The combined Agostino K^2^ test was significant for the CHRT-PR_16_ model (*p* < .001) and borderline non-significant for the CHRT-PR_14_ model (*p* = .06), indicating that the CHRT-PR_14_ shows somewhat better overall residual calibration than the CHRT-PR_16_, though neither fully resolves the kurtosis limitation.

### Model Validation and Normative Trajectories

The final ZINBI models demonstrated excellent empirical calibration. In the 80/20 train-test holdout validation, the empirical proportions falling at or below the model-derived 50^th^ percentile aligned closely with the expected .50 threshold. For the CHRT-PR_16_, the held-out calibration was .525 for males (n = 242) and .546 for females (n = 205). For the CHRT-PR_14_, held out calibration was .525 for males and .522 for females.

Calculated age curves and final lookup tables revealed systematic sex and age divergence (Table 3 and Figures 2 and 3). Males generally exhibited slightly higher total scores at comparable age and percentile thresholds that began around age 10 and mostly continued through age 17. This divergence resulted in systematic 1- to 2-point shifts in percentile cutoff scores between sexes.

**Figure 2.**
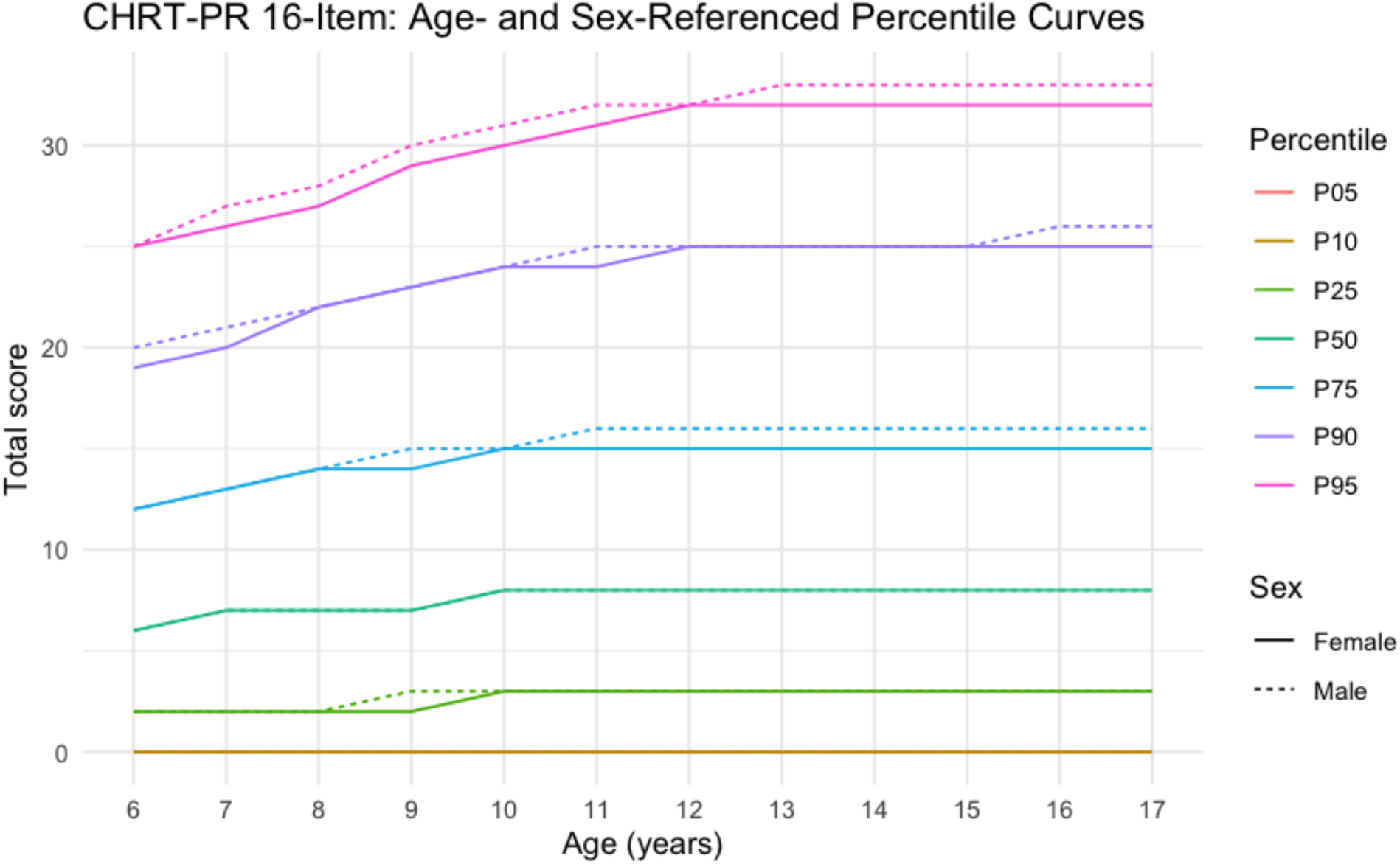
Continuous Age- and Sex-Referenced Centile Growth Curves for the CHRT-PR_16_. *Note.* Continuous centile growth curves modeled via Generalized Additive Models for Location, Scale and Shape (GAMLSS) under a Zero-Inflated Negative Binomial (ZINBI) distribution for the CHRT-PR_16_ across youth aged 6 to 18 (*N* = 2,235; males *n* = 1,123; females *n* = 1,112). Curves depict the 10^th^, 25^th^, 50^th^, 75^th^, 90^th^, and 95^th^ population percentiles across age for males and females. Trajectories diverge by sex starting around age 10, with males showing higher scores at upper percentiles in early adolescence. *Abbreviations:* GAMLSS = Generalized Additive Models for Location, Scale and Shape; ZINBI = Zero-Inflated Negative Binomial.

**Figure 3.**
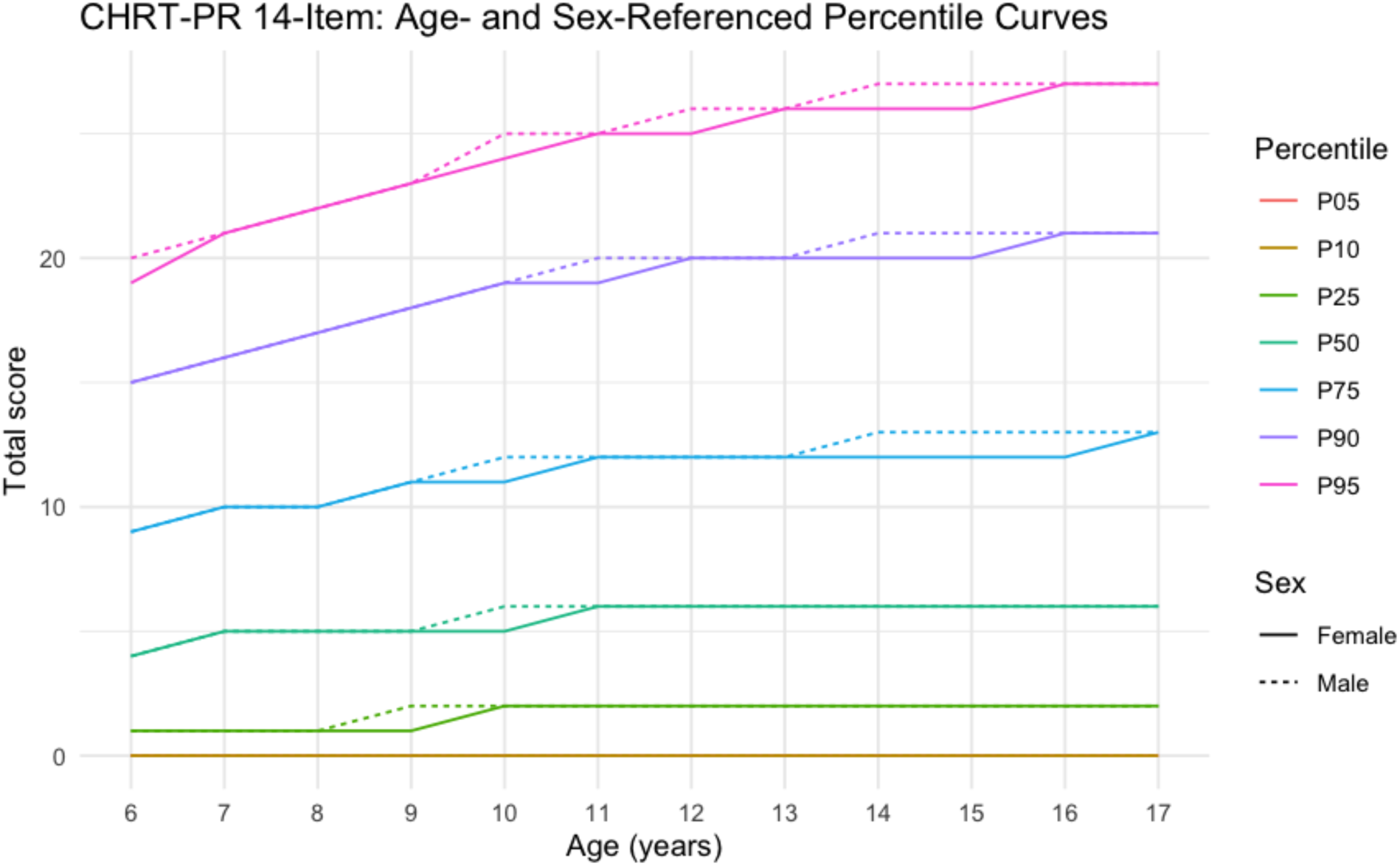
Continuous Age- and Sex-Referenced Centile Growth Curves for the CHRT-PR_14_. *Note.* Continuous centile growth curves modeled via Generalized Additive Models for Location, Scale and Shape (GAMLSS) under a Zero-Inflated Negative Binomial (ZINBI) distribution for the CHRT-PR_14_ across youth aged 6 to 18 (*N* = 2,235; males *n* = 1,123; females *n* = 1,112). Curves depict the 10^th^, 25^th^, 50^th^, 75^th^, 90^th^, and 95^th^ population percentiles across age for males and females. Trajectories diverge by sex starting around age 10, with males showing higher scores at upper percentiles in early adolescence. *Abbreviations:* GAMLSS = Generalized Additive Models for Location, Scale and Shape; ZINBI = Zero-Inflated Negative Binomial.

**Table 3.** Continuous Age- and Sex-Referenced Percentile Cutoff Scores for CHRT-PR_16_ and CHRT-PR_14_.

| **Age**<br>*(Years)* | **25th %ile**<br>*Male / Female* | **50th %ile**<br>*Male / Female* | **75th %ile**<br>*Male / Female* | **90th %ile**<br>*Male / Female* | **95th %ile**<br>*Male / Female* |
| --- | --- | --- | --- | --- | --- |
| ***CHRT-PR<sub>16</sub> Percentile Cutoffs*** |  |  |  |  |  |
| 6 | 2 / 2 | 6 / 6 | 12 / 12 | 20 / 19 | 25 / 25 |
| 7 | 2 / 2 | 7 / 7 | 13 / 13 | 21 / 20 | 27 / 26 |
| 8 | 2 / 2 | 7 / 7 | 14 / 14 | 22 / 22 | 28 / 28 |
| 9 | 3 / 2 | 7 / 7 | 15 / 14 | 23 / 23 | 30 / 29 |
| 10 | 3 / 3 | 8 / 8 | 15 / 15 | 24 / 24 | 31 / 30 |
| 11 | 3 / 3 | 8 / 8 | 16 / 15 | 25 / 24 | 32 / 31 |
| 12 | 3 / 3 | 8 / 8 | 16 / 15 | 25 / 25 | 32 / 32 |
| 13 | 3 / 3 | 8 / 8 | 16 / 15 | 25 / 25 | 32 / 32 |
| 14 | 3 / 3 | 8 / 8 | 16 / 15 | 25 / 25 | 32 / 32 |
| 15 | 3 / 3 | 8 / 8 | 16 / 15 | 25 / 25 | 32 / 32 |
| 16 | 3 / 3 | 8 / 8 | 16 / 15 | 25 / 25 | 33 / 32 |
| 17 | 3 / 3 | 8 / 8 | 16 / 15 | 26 / 25 | 33 / 32 |
| 18 | 3 / 3 | 8 / 8 | 16 / 15 | 26 / 25 | 33 / 32 |
| ***CHRT-PR<sub>14</sub> Percentile Cutoffs*** |  |  |  |  |  |
| 6 | 1 / 1 | 4 / 4 | 9 / 9 | 15 / 15 | 20 / 20 |
| 7 | 1 / 1 | 5 / 5 | 10 / 10 | 16 / 16 | 21 / 21 |
| 8 | 1 / 1 | 5 / 5 | 10 / 10 | 17 / 17 | 22 / 22 |
| 9 | 2 / 1 | 5 / 5 | 11 / 11 | 18 / 18 | 23 / 23 |
| 10 | 2 / 2 | 6 / 5 | 12 / 11 | 19 / 19 | 25 / 24 |
| 11 | 2 / 2 | 6 / 6 | 12 / 12 | 20 / 19 | 25 / 25 |
| 12 | 2 / 2 | 6 / 6 | 12 / 12 | 20 / 20 | 26 / 25 |
| 13 | 2 / 2 | 6 / 6 | 12 / 12 | 20 / 20 | 26 / 26 |
| 14 | 2 / 2 | 6 / 6 | 13 / 12 | 21 / 20 | 27 / 26 |
| 15 | 2 / 2 | 6 / 6 | 13 / 12 | 21 / 20 | 27 / 26 |
| 16 | 2 / 2 | 6 / 6 | 13 / 12 | 21 / 21 | 27 / 27 |
| 17 | 2 / 2 | 6 / 6 | 13 / 13 | 21 / 21 | 27 / 27 |
| 18 | 2 / 2 | 6 / 6 | 13 / 13 | 21 / 21 | 28 / 27 |

*Note.* Percentile cutoffs derived from Zero-Inflated Negative Binomial (ZINBI) Generalized Additive Models for Location, Scale and Shape (GAMLSS). Values represent unit-weighted raw sum scores.

**Table 4.**
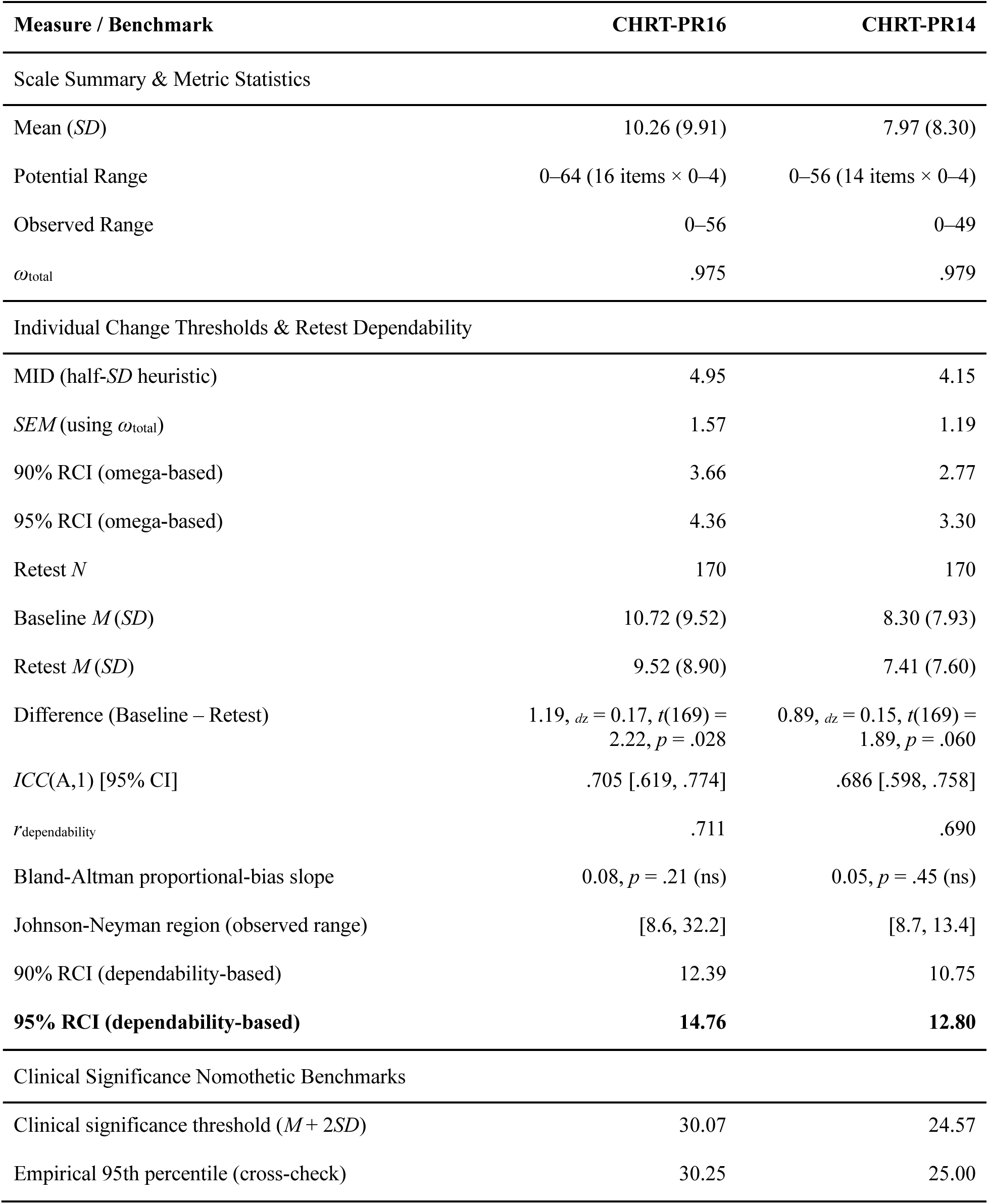
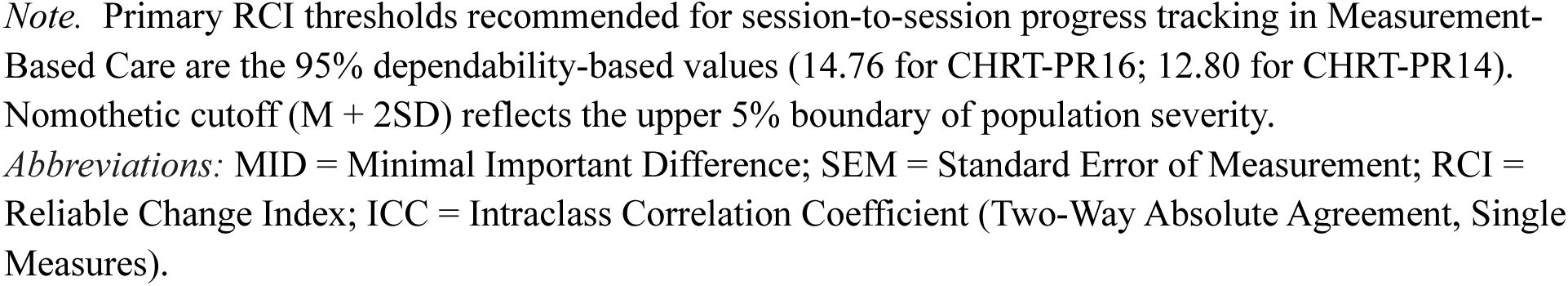
Clinical Significance and Reliable Change Benchmarks for CHRT-PR16 and CHRT-PR14.

| Measure / Benchmark | CHRT-PR16 | CHRT-PR14 |
| --- | --- | --- |
| Scale Summary & Metric Statistics |  |  |
| Mean (*SD*) | 10.26 (9.91) | 7.97 (8.30) |
| Potential Range | 0–64 (16 items × 0–4) | 0–56 (14 items × 0–4) |
| Observed Range | 0–56 | 0–49 |
| $\omega_{\text{total}}$ | .975 | .979 |
| Individual Change Thresholds & Retest Dependability |  |  |
| MID (half-*SD* heuristic) | 4.95 | 4.15 |
| *SEM* (using $\omega_{\text{total}}$) | 1.57 | 1.19 |
| 90% RCI (omega-based) | 3.66 | 2.77 |
| 95% RCI (omega-based) | 4.36 | 3.30 |
| Retest *N* | 170 | 170 |
| Baseline *M* (*SD*) | 10.72 (9.52) | 8.30 (7.93) |
| Retest *M* (*SD*) | 9.52 (8.90) | 7.41 (7.60) |
| Difference (Baseline – Retest) | 1.19, $d_z = 0.17$, $t(169) = 2.22$, $p = .028$ | 0.89, $d_z = 0.15$, $t(169) = 1.89$, $p = .060$ |
| *ICC*(A,1) [95% CI] | .705 [.619, .774] | .686 [.598, .758] |
| $r^{\text{dependability}}$ | .711 | .690 |
| Bland-Altman proportional-bias slope | 0.08, $p = .21$ (ns) | 0.05, $p = .45$ (ns) |
| Johnson-Neyman region (observed range) | [8.6, 32.2] | [8.7, 13.4] |
| 90% RCI (dependability-based) | 12.39 | 10.75 |
| **95% RCI (dependability-based)** | **14.76** | **12.80** |
| Clinical Significance Nomothetic Benchmarks |  |  |
| Clinical significance threshold ($M + 2SD$) | 30.07 | 24.57 |
| Empirical 95th percentile (cross-check) | 30.25 | 25.00 |

*Note.* Primary RCI thresholds recommended for session-to-session progress tracking in Measurement-Based Care are the 95% dependability-based values (14.76 for CHRT-PR16; 12.80 for CHRT-PR14). Normothetic cutoff ( $M + 2SD$ ) reflects the upper 5% boundary of population severity.
*Abbreviations:* MID = Minimal Important Difference; SEM = Standard Error of Measurement; RCI = Reliable Change Index; ICC = Intraclass Correlation Coefficient (Two-Way Absolute Agreement, Single Measures).

### Clinical Significance Benchmarks

Beyond statistical reliability, two benchmarks were derived to support interpretation of individual scores and score changes in clinical terms. The MID, estimated using the conventional half-*SD* heuristic (Norman et al., 2003), was 4.95 points for CHRT-PR_16_ and 4.15 points for CHRT-PR_14_. Reliable change was benchmarked using the 2-week retest subsample, reflecting genuine short-term symptom fluctuation rather than only item-sampling error. Test-retest correlations were .71 (CHRT-PR_16_) and .69 (CHRT-PR_14_). A change exceeding 14.8 points (CHRT-PR_16_) or 12.8 points (CHRT-PR_14_) exceeds measurement and short-term state variability with 95% confidence. This is a substantially higher bar than an internal-consistency-only estimate would suggest, and the more defensible threshold for judging whether an individual’s change over time reflects genuine clinical improvement or deterioration.

A second benchmark evaluates extreme severity relative to population distributions. Following Jacobson and Truax’s (1991) widely used framework, one of the three cutoff definitions that framework offers, only one can be computed from a general-population norming sample alone and is defined as a score falling within two standard deviations of the normative (nonclinical) population mean. Here, this benchmark is a score of 30.07 points for CHRT-PR_16_ and 24.57 points for CHRT-PR_14_, both closely matching this sample’s own empirical 95th percentile (30.25 and 25.00, respectively). For high-severity youth presenting with acute distress, moving below this benchmark may represent a critical initial treatment milestone as it marks a patient’s transition out of the extreme tail of population risk as care proceeds toward full remission.

To further characterize agreement between the baseline and retest administrations, intraclass correlation, raw score difference, and Bland-Altman analyses were conducted using the same strict-matched retest subsample. Mean scores were 10.72 (*SD* = 9.52) at baseline and 9.52 (*SD* = 8.90) at retest for CHRT-PR_16_, and 8.30 (*SD* = 7.93) at baseline and 7.41 (*SD* = 7.60) at retest for CHRT-PR_14_. Absolute-agreement ICCs, ICC(A,1), were .70 (CHRT-PR_16_) and .69 (CHRT-PR_14_). Scores decreased slightly from baseline to retest for both scales (CHRT-PR_16_: *M_diff* = 1.19 points, *d_z* = 0.17, *t*(169) = 2.22, *p* = .028; CHRT-PR_14_: *M_diff* = 0.89, *d_z =* 0.15, *t*(169) = 1.89, *p* = .060), consistent with regression to the mean and/or practice effects rather than genuine symptom change, since no intervention occurred between administrations. Bland-Altman analysis found no evidence of proportional bias for either scale (CHRT-PR_16_: *b* = 0.08, *p* = .21; CHRT-PR_14_: *b* = 0.05, *p* = .45), indicating the magnitude of the baseline-retest difference did not vary systematically with overall symptom severity (see Supplemental Figures S3 and S4). A Johnson-Neyman analysis identified a region of each scale’s observed score range within which the average difference was statistically distinguishable from zero (CHRT-PR_16_: mean scores 8.6-32.2; CHRT-PR_14_: 8.7-13.4). Because this region falls in the middle of the observed range rather than at either extreme and the proportional-bias slope was flat, it reflects the greater precision of the estimate where the retest sample is densest, not evidence that the difference itself varies by severity.

## Discussion

Establishing national norms for parent-reported pediatric suicide risk fills a vital gap in measurement-based care, providing clinicians with population-level reference points to contextualize symptom severity, track treatment response, and detect meaningful clinical change (Hua et al., 2024; Weitzman et al., 2025; Mayes et al., 2020). We found that both item sets behave as “essentially unidimensional.” When more than one factor was present, the variance attributable to the general factor dominated, and the variance in smaller group factors was not enough to justify forming subscales on a psychometric basis, let alone in terms of clinical utility. All items showed moderate to excellent discrimination and factor loadings, and the reliability of the scores was high across a broad range of severity. The raw total and IRT-based scoring correlated so highly that there was no practical advantage in using IRT scoring. There was no evidence of differential item functioning across sex or three age strata. Overall, the use of a total item score shows excellent performance in a general population sample as opposed to more complex scoring approaches.

In addition to creating and validating the parent-report CHRT-PR_16_ and CHRT-PR_14_, we provide age- and sex-specific percentile lookup tables for the CHRT-PR_16_ and CHRT-PR_14_. Additionally, score trajectories demonstrate age and sex divergence beginning around age 10, producing 1- to 2-point shifts in cutoff thresholds and highlighting the need for sex-specific normative tables in pediatric settings. Both the CHRT-PR_16_ and CHRT-PR_14_ were well-supported as single-score measures of overall suicide risk, each dominated by one general factor accounting for the large majority of reliable variance (ω_H_ = .68 and .83, respectively). Neither version is clearly superior: CHRT-PR_14_ shows modestly cleaner item behavior and a better-fitting norming model, while CHRT-PR_16_ shows consistently stronger, albeit marginally, correlations with external criteria spanning depression, functional impairment, and service utilization. Taken together, the choice between CHRT-PR_16_ and CHRT-PR_14_ depends on practical issues such as respondent burden and whether the two additional irritability items add clinically actionable information for a given use case as opposed to psychometric issues.

To account for the severe floor effects and right-skewness of general population screening data for suicide risk, GAMLSS ZINBI regression was used to derive continuous norms, achieving optimal fit (BIC) across both CHRT-PR_16_ and CHRT-PR_14_. GAMLSS modeling highlighted that both the CHRT-PR_16_ and CHRT-PR_14_ fully resolved residual skewness, although a residual kurtosis limitation remained for both versions, which was most pronounced in early-to-mid adolescent age bands, and otherwise provided stable reference norms. Local dependence was concentrated within theoretically related item pairs (e.g., irritability and impulsivity), suggesting clinically meaningful content overlap rather than widespread model misspecification.

Although approximately 20% of item pairs exceeded the conventional threshold, misfit was localized to a small number of conceptually similar item clusters, and total scores remained highly consistent with IRT-based trait estimates. Interestingly, our cutoff scores for the 90^th^ and 95^th^ percentiles for both males and females on the CHRT-PR_14_ were very closely aligned to the optimal cutoff scores for predicting a suicide attempt provided by Mayes et al., 2020, at least for older adolescents. Specifically, Mayes et al., (2020) found that a cutoff score of 28 predicted a suicide attempt (85.7% sensitivity, 56.5% specificity) and a score of 22 predicted a suicidal event (defined as an attempt or an emergency department visit or hospitalization due to worsening suicidal ideation; 80.8% sensitivity, 40.9% specificity) in their adolescent sample. For adolescents 16-17 years old, our 90^th^ and 95^th^ percentile cutoffs for the CHRT-PR_14_ were 21 and 27 respectively, demonstrating strong alignment with established clinical risk thresholds.

While our growth chart shows that males and females show relatively similar cutoff scores across development, sex differences in cutoff scores are evident at the higher thresholds. Despite only 1-to 2-point shifts between males and females, use of sex-specific reference tables is indicated to reduce error in over-identifying risk in males or under-identifying risk in females. This divergence begins around age 10, in which males frequently have higher cutoff scores relative to females beyond this point. Importantly, multigroup CFA and DIF analyses confirmed full scalar invariance across sex and age strata for both CHRT-PR_16_ and CHRT-PR_14_, establishing that both measure suicide risk with equal item fairness across demographic groups. Consequently, the score divergence observed after age 10 is unlikely to be attributable to measurement artifact, suggesting that the observed differences may reflect developmental variation and providing empirical justification for sex-specific normative lookup tables. While males are more likely to die by suicide relative to females, females are more likely to report suicidal ideation and behavior, which makes this discrepancy opposite of what would be anticipated (Hua et al., 2024; Carretta et al., 2023; Xiao et al., 2021). This could be an artifact of informant reporting, in which males may demonstrate more observable suicide risk indicators in adolescence. However, prior research demonstrates that parental awareness of suicide risk in males *decreases* in adolescence, while parental awareness for females increases (Jones et al., 2019). Future analyses examining our youth report data will help elucidate this discrepancy.

Anchoring the RCI to short-term test-retest dependability yields a conservative 95% RCI threshold of 14.8 points (CHRT-PR_16_) and 12.8 points (CHRT-PR_14_). This provides a benchmark for session-to-session progress tracking. In the 2-week retest subsample, raw scores exhibited a small natural decrease from baseline, reflecting expected regression to the mean and practice effects in the absence of known treatment. This routine score fluctuation fell far below both the MID and the RCI threshold. This pattern demonstrates the practical utility of the RCI in MBC given that clinicians can rely on the reliable change threshold rather than raw score decreases. However, this drift should nonetheless be read as a floor rather than a universal estimate given that this is a normative sample. Specifically, this sample’s score range does not extend to the more elevated severity levels typical of a treatment-seeking population, so whether the same flat pattern holds at higher severity (where regression to the mean would be expected to be larger) remains an open question this dataset cannot resolve.

We also established benchmark scores of 30.07 and 24.57 for CHRT-PR_16_ and CHRT-PR_14_ per Jacobson and Truax’s framework (1991). These cutoffs approximate the 95^th^ percentile of scores, indicating concerning elevations. However, this benchmark may provide a valuable initial treatment target when working with youth presenting with extremely high baseline scores, marking a patient’s de-escalation out of the extreme tail of population risk. Future longitudinal validation in treatment-seeking clinical cohorts will need to build upon these normative benchmarks to evaluate clinical recovery boundaries against prospective suicidal behavior and acute care utilization.

The present study has several strengths and weaknesses. While the community sample is ideal for establishing non-clinical normative benchmarks, future research is needed to evaluate these cutoffs in clinical samples. The current study emphasized convergent validity and did not examine discriminant validity relative to constructs theoretically distinct from suicide risk. Future work should evaluate the extent to which CHRT-PR_16_ and CHRT-PR_14_ scores capture variance beyond broader psychopathology and general distress. Further, the use of parent report is limited to capturing observable behavioral manifestations of risk, which may differ from self-reported internal distress (Lewis et al., 2014). Subsequent analyses with BENCHMARK data will establish norms using youth report data and evaluate discrepancies between parent and youth report as multi-informant assessment remains the gold standard in youth behavioral health. This is especially true for younger children. Future longitudinal studies are needed to evaluate how these normative cutoffs predict prospective suicidal behavior across diverse clinical cohorts.

Overall, the present study establishes the first national normative tables to assist clinicians in contextualizing parent-reported suicide risk against a nationally representative population, facilitating tracking progress across treatment and identifying meaningful risk improvement or deterioration relative peers of the same age and sex. This work provides compelling psychometric and distributional support for the use of either the CHRT-PR_16_ or CHRT-PR_14_ in clinical settings.

## Acknowledgements

This work was supported by gifts from the Chlapaty, DiMarco, and Zipfel families to Nationwide Children’s Hospital for the Institute for Mental and Behavioral Health Research (IMBHR). The authors gratefully acknowledge Marissa McClellan, Yinuo Liu, Ellie Nienaber, Kevin Stephenson, Jacqui Pazarapolous, Brittany Grate, and Kim Jones for their contributions to the development and implementation of the BENCHMARK project, including literature and reference review, study coordination and administration, data organization and management, data cleaning, and development of analytic resources.

## Declaration of Interest Statement

Dr. Youngstrom is the co-founder and Executive Director of Helping Give Away Psychological Science, a 501c3; he receives funding from the National Institutes of Health, and he has received royalties from the American Psychological Association and Guilford Press, and he holds equity in Joe Startup Technologies. Dr. Westlund Schreiner receives funding from the Brain and Behavior Research Foundation and the National Institutes of Health.

## CRediT roles

EAY conceptualized the project and secured provision of nationally representative data; HD, EAY, and MWS ran analyses and created visualizations; HD and MWS wrote all sections of the original draft; MWS and EAY provided supervision; all authors reviewed and edited the full manuscript.

## Funding Details

No intramural funding was obtained for the reported work.

## Declaration of AI Use

Generative AI [ChatGPT Model 5.6 Sol] was used to 1) support code debugging in R, with all code checked and run by authors, Claude Code was used for an independent reanalysis and verification of the reproducibility of results; 2) copyedit the final manuscript draft; and 3) review adherence to standard BENCHMARK study procedures. An OpenEvidence search was used to look for any additional references pertaining to CHRT measurement in youths.

## Data Availability Statement

Code and sufficient data for replication available upon request from the corresponding author.

## Notes

### Author Declarations

This study was approved by the Institutional Review Board at Nationwide Childrens Hospital.

## References

Achenbach, T. M. (1995). Empirically based assessment and taxonomy: Applications to clinical research. Psychological Assessment, 7(3), 261–274. 10.1037/1040-3590.7.3.261

Achenbach, T. M. (2001). What are norms and why do we need valid ones?

Achenbach, T. M., & Rescorla, L. A. (2001). Child Behavior Checklist for Ages 6-18. University of Vermont Burlington, VT.

Batterham, P. J., Ftanou, M., Pirkis, J., Brewer, J. L., Mackinnon, A. J., Beautrais, A., Fairweather-Schmidt, A. K., & Christensen, H. (2015). A systematic review and evaluation of measures for suicidal ideation and behaviors in population-based research. Psychological Assessment, 27(2), 501–512. 10.1037/pas0000053

Carretta, R. F., McKee, S. A., & Rhee, T. G. (2023). Gender Differences in Risks of Suicide and Suicidal Behaviors in the USA: A Narrative Review. Current Psychiatry Reports, 25(12), 809–824. 10.1007/s11920-023-01473-1

Centers for Disease Control and Prevention. (2024). Web-based Injury Statistics Query and Reporting System (WISQARS).

Chen, F. F. (2007). Sensitivity of goodness of fit indexes to lack of measurement invariance. Structural Equation Modeling, 14(3), 464–504. 10.1080/10705510701301834

Cheung, G. W., & Rensvold, R. B. (2002). Evaluating goodness-of-fit indexes for testing measurement invariance. Structural Equation Modeling, 9(2), 233–255. 10.1207/S15328007SEM0902_5

Choi, S. W., Gibbons, L. E., & Crane, P. K. (2009). lordif: Logistic Ordinal Regression Differential Item Functioning using IRT (p. 0.4.2) [Dataset]. 10.32614/CRAN.package.lordif

Covarrubias, J. J., & Fristad, M. A. (2025). A Brief Quality of Life (QoL) Scale: Initial Psychometric Data for the Nationwide QoL Scale (NQLS). Journal of Psychopathology and Behavioral Assessment, 47(3), 66. 10.1007/s10862-025-10244-6

Cronbach, L. J. (1951). Coefficient alpha and the internal structure of tests. Psychometrika, 16(3), 297–334. 10.1007/BF02310555

De La Garza, N., John Rush, A., Grannemann, B. D., & Trivedi, M. H. (2017). Toward a very brief self-report to assess the core symptoms of depression (VQIDS-SR_5_ ). Acta Psychiatrica Scandinavica, 135(6), 548–553. 10.1111/acps.12720

De Los Reyes, A., & Epkins, C. C. (2023). Introduction to the Special Issue. A Dozen Years of Demonstrating That Informant Discrepancies are More Than Measurement Error: Toward Guidelines for Integrating Data from Multi-Informant Assessments of Youth Mental Health. Journal of Clinical Child & Adolescent Psychology, 52(1), 1–18. 10.1080/15374416.2022.2158843

Drasgow, F., Levine, M. V., & Williams, E. A. (1985). Appropriateness measurement with polychotomous item response models and standardized indices. British Journal of Mathematical and Statistical Psychology, 38(1), 67–86. 10.1111/j.2044-8317.1985.tb00817.x

Feuerstahler, L. M., Waller, N., & MacDonald, A. (2020). Improving Measurement Precision in Experimental Psychopathology Using Item Response Theory. Educational and Psychological Measurement, 80(4), 695–725. 10.1177/0013164419892049

Gadow, K. D. & S. (2016). Child & Adolescent Symptom Inventory-5. Checkmate Plus.

Gardner, W., Murphy, M., Childs, G., Kelleher, K., Pagano, M., Jellinek, M., McInerny, T. K., Wasserman, R. C., Nutting, P., Chiappetta, L., & Sturner, R. (1999). The PSC-17: A brief pediatric symptom checklist with psychosocial problem subscales. A report from PROS and ASPN. Ambulatory Child Health, *5*(3), 225–236.

Hua, L. L., Lee, J., Rahmandar, M. H., Sigel, E. J., COMMITTEE ON ADOLESCENCE, & COUNCIL ON INJURY, VIOLENCE, AND POISON PREVENTION. (2024). Suicide and Suicide Risk in Adolescents. Pediatrics, 153(1), e2023064800. 10.1542/peds.2023-064800

Jacobson, N. S., & Truax, P. (1991). Clinical significance: A statistical approach to defining meaningful change in psychotherapy research. Journal of Consulting and Clinical Psychology, 59(1), 12–19. 10.1037/0022-006X.59.1.12

Jones, J. D., Boyd, R. C., Calkins, M. E., Ahmed, A., Moore, T. M., Barzilay, R., Benton, T. D., & Gur, R. E. (2019). Parent-Adolescent Agreement About Adolescents’ Suicidal Thoughts. Pediatrics, 143(2), e20181771. 10.1542/peds.2018-1771

Lewis, A. J., Bertino, M. D., Bailey, C. M., Skewes, J., Lubman, D. I., & Toumbourou, J. W. (2014). Depression and suicidal behavior in adolescents: A multi-informant and multi-methods approach to diagnostic classification. Frontiers in Psychology, 5. 10.3389/fpsyg.2014.00766

Mayes, T. L., Carmody, T., Rush, A. J., Nandy, K., Emslie, G. J., Kennard, B. D., Forbes, K., Jha, M. K., Hughes, J. L., Heerschap, J. K., & Trivedi, M. H. (2023). Predicting suicidal events: A comparison of the Concise Health Risk Tracking Self-Report (CHRT-SR) and the Columbia Suicide Severity Rating Scale (C-SSRS). Psychiatry Research, 326, 115306. 10.1016/j.psychres.2023.115306

Mayes, T. L., Killian, M., Rush, A. J., Emslie, G. J., Carmody, T., Kennard, B. D., Jha, M. K., King, J., Hughes, J. L., & Trivedi, M. H. (2020). Predicting future suicidal events in adolescents using the Concise Health Risk Tracking Self-Report (CHRT-SR). Journal of Psychiatric Research, 126, 19–25. 10.1016/j.jpsychires.2020.04.008

McClellan, M. B., Rush, A. J., & Youngstrom, E. A. (2025). *Nationwide Gauge of Emotional Severity (N-Gauge)*. Institute for Mental and Behavioral Health Research, Nationwide Children’s Hospital.

McDonald, R. P. (1999). Test theory: A unified treatment (pp. xi, 485). Lawrence Erlbaum Associates Publishers.

Norman, G. R., Sloan, J. A., & Wyrwich, K. W. (2003). Interpretation of Changes in Health-related Quality of Life: The Remarkable Universality of Half a Standard Deviation. Medical Care, 41(5), 582–592. 10.1097/01.MLR.0000062554.74615.4C

Ostacher, M. J., Nierenberg, A. A., Rabideau, D., Reilly-Harrington, N. A., Sylvia, L. G., Gold, A. K., Shesler, L. W., Ketter, T. A., Bowden, C. L., Calabrese, J. R., Friedman, E. S., Iosifescu, D. V., Thase, M. E., Leon, A. C., & Trivedi, M. H. (2015). A clinical measure of suicidal ideation, suicidal behavior, and associated symptoms in bipolar disorder: Psychometric properties of the Concise Health Risk Tracking Self-Report (CHRT-SR). Journal of Psychiatric Research, 71, 126–133. 10.1016/j.jpsychires.2015.10.004

Posner, K., Brown, G. K., Stanley, B., Brent, D. A., Yershova, K. V., Oquendo, M. A., Currier, G. W., Melvin, G. A., Greenhill, L., Shen, S., & Mann, J. J. (2011). The Columbia–Suicide Severity Rating Scale: Initial Validity and Internal Consistency Findings From Three Multisite Studies With Adolescents and Adults. American Journal of Psychiatry, 168(12), 1266–1277. 10.1176/appi.ajp.2011.10111704

Rosseel, Y. (2012). lavaan: An R Package for Structural Equation Modeling. Journal of Statistical Software, 48, 1–36. 10.18637/jss.v048.i02

Sprafkin, J., Gadow, K. D., Salisbury, H., Schneider, J., & Loney, J. (2002). Further evidence of reliability and validity of the Child Symptom Inventory-4: Parent checklist in clinically referred boys. Journal of Clinical Child & Adolescent Psychology, 31(4), 513–524. 10.1207/S15374424JCCP3104_10

Timmerman, M. E., De Bildt, A., & Urban, J. (2026). The GRoNC: Guidelines for Reporting on Norm-Referenced and Criterion-Referenced Scores. Assessment, 33(6), 954–972. 10.1177/10731911251371395

Timmerman, M. E., Voncken, L., & Albers, C. J. (2021). A tutorial on regression-based norming of psychological tests with GAMLSS. Psychological Methods, 26(3), 357–373. 10.1037/met0000348

Trivedi, M. H., Wisniewski, S. R., Morris, D. W., Fava, M., Gollan, J. K., Warden, D., Nierenberg, A. A., Gaynes, B. N., Husain, M. M., Luther, J. F., Zisook, S., & Rush, A. J. (2011). Concise Health Risk Tracking Scale: A Brief Self-Report and Clinician Rating of Suicidal Risk. The Journal of Clinical Psychiatry, 72(06), 757–764. 10.4088/JCP.11m06837

Trombello, J. M., Kulikova, A., Mayes, T. L., Nandy, K., Carmody, T., Bart, G., Nunes, E. V., Schmitz, J., Kalmin, M., Shoptaw, S., & Trivedi, M. H. (2023). Psychometrics of the Concise Health Risk Tracking Self-Report (CHRT-SR16) Assessment of Suicidality in a Sample of Adults with Moderate to Severe Methamphetamine Use Disorder: Findings from the ADAPT-2 Randomized Trial. Neuropsychiatric Disease and Treatment, *Volume 19*, 1443–1454. 10.2147/NDT.S406909

Weitzman, C., Guevara, J., Curtin, M., Macias, M., AAP Section on Developmental and Behavioral Pediatrics, Kinwa Poon, J., Joseph Smith, P. J., Christine Augustyn, M., Liu, Y. H., Anopawuia Spinks-Franklin, A. I., Zubler, J. M., AAP Council on Early Childhood, Navsaria, D., Glusman, M., Uzoatu Anyigbo, C., Chen, V., Gonzales, J. L., Guevara, J. P., Nobuhide Hashikawa, A., … Degnon, L. (2025). Promoting Optimal Development: Screening for Mental Health, Emotional, and Behavioral Problems: Clinical Report. Pediatrics, *156*(3), e2025073172. 10.1542/peds.2025-073172

Xiao, Y., Cerel, J., & Mann, J. J. (2021). Temporal Trends in Suicidal Ideation and Attempts Among US Adolescents by Sex and Race/Ethnicity, 1991-2019. JAMA Network Open, *4*(6), e2113513. 10.1001/jamanetworkopen.2021.13513

Yamamoto, K., Khorramdel, L., & von Davier, M. (2013). Scaling PIAAC cognitive data. OECD, Technical Report of the Survey of Adult Skills (PIAAC) (Ch. 17).

Yen, W. M. (1984). Effects of Local Item Dependence on the Fit and Equating Performance of the Three-Parameter Logistic Model. Applied Psychological Measurement, 8(2), 125–145. 10.1177/014662168400800201

Youngstrom, E. A., Youngstrom, J. K., Freeman, A. J., De Los Reyes, A., Feeny, N. C., & Findling, R. L. (2011). Informants Are Not All Equal: Predictors and Correlates of Clinician Judgments About Caregiver and Youth Credibility. Journal of Child and Adolescent Psychopharmacology, 21(5), 407–415. 10.1089/cap.2011.0032

