## Supplement for "Psychometric Properties and Continuous National Norms of the Parent-Report Concise Health Risk Tracking Assessment: Charting Pediatric Suicide Risk Across Age and Sex"

**Supplemental Table S1**

*CHRT-PR$_{16}$ GRM item parameters*

| Item | a | b1 | b2 | b3 | b4 | Discrimination |
|---|---|---|---|---|---|---|
| Never going to get better (1) | 2.74 | 0.14 | 1.01 | 1.70 | 2.57 | very high |
| No future (2) | 3.76 | 0.57 | 1.33 | 1.88 | 2.53 | very high |
| Can do nothing right (3) | 2.81 | 0.21 | 1.07 | 1.69 | 2.50 | very high |
| Everything turns out wrong (4) | 3.07 | 0.23 | 1.14 | 1.68 | 2.67 | very high |
| No one to depend on (5) | 3.91 | 0.63 | 1.42 | 1.92 | 2.73 | very high |
| People cared for are gone (6) | 2.55 | 0.68 | 1.52 | 2.08 | 2.66 | very high |
| Wishes suffering were over (7) | 3.25 | 0.74 | 1.31 | 1.83 | 2.51 | very high |
| No reason to live (8) | 6.04 | 0.93 | 1.51 | 1.96 | 2.63 | very high |
| Wishes to sleep, not wake (9) | 4.94 | 0.97 | 1.56 | 1.93 | 2.45 | very high |
| Acts without thinking (10) | 1.39 | −0.45 | 0.39 | 1.17 | 2.93 | high |
| Decisions on impulse (11) | 1.25 | −0.76 | 0.24 | 1.12 | 2.99 | moderate |
| *Irritable/easily angered (12)* | 1.71 | −0.36 | 0.57 | 1.22 | 2.48 | very high |
| *Overreacts with anger/rage (13)* | 1.69 | −0.11 | 0.79 | 1.34 | 2.49 | high |
| Thoughts of killing self (14) | 5.43 | 1.01 | 1.52 | 1.85 | 2.53 | very high |
| Thoughts how to kill self (15) | 5.06 | 1.03 | 1.55 | 1.98 | 2.41 | very high |
| Has a plan to kill self (16) | 4.42 | 1.17 | 1.71 | 2.26 | 2.72 | very high |

*Note.* Item parameters estimated using a unidimensional Graded Response Model (GRM) on the complete-case baseline sample ($N = 2{,}236$). Parameter $a$ represents item discrimination (slopes), categorized according to Baker and Kim’s (2017) criteria (moderate = 0.65–1.34; high = 1.35–1.69; very high ≥ 1.70). Parameters $b1$ through $b4$ represent item difficulty thresholds (category boundary locations along the latent trait θ scale). Italicized items (12 and 13) are excluded from the CHRT-PR14 short form. *Abbreviations:* GRM = Graded Response Model; CHRT-PR$_{16}$ = Concise Health Risk Tracking 16-Item Parent Report.

**Supplemental Table S2**

*Item Fit RMSD for CHRT-PR$_{16}$*

| Item | RMSD |
|---|---|
| Decisions on impulse (11) | .058 |
| Acts without thinking (10) | .050 |
| Irritable/easily angered (12) | .050 |
| Overreacts with anger/rage (13) | .047 |
| Everything turns out wrong (4) | .028 |
| Can do nothing right (3) | .027 |
| Never going to get better (1) | .018 |
| Wishes suffering were over (7) | .016 |
| No future (2) | .013 |
| People cared for are gone (6) | .013 |
| No one to depend on (5) | .012 |
| Thoughts how to kill self (15) | .011 |
| Has a plan to kill self (16) | .008 |
| No reason to live (8) | .007 |
| Wishes to sleep, not wake (9) | .007 |
| Thoughts of killing self (14) | .007 |

*Note.* Item-level Root Mean Square Deviation (RMSD) absolute model fit statistics computed manually per Yamamoto et al. (2013) across decile bins of EAP θ trait estimates. Values of RMSD ≤ .10 indicate acceptable item-level model fit. Zero items across either scale length exceeded the .10 misfit boundary. *Abbreviations:* EAP = Expected A Posteriori; GRM = Graded Response Model; RMSD = Root Mean Square Deviation.

**Supplemental Table S3**

*Summary of GRM Item and Person Fit Diagnostics*

| Scale | Items | Items flagged (RMSD > .10) | Mean $Q_3$ | Pairs flagged ($\|Q_3\| > .20$) | $N$ | Persons flagged ($\|Z_h\| > 3$) |
|---|---|---|---|---|---|---|
| CHRT-PR16 | 16 | 0 (0%) | -.045 | 25/120 (20.8%) | 2236 | 35 (1.57%) |
| CHRT-PR14 | 14 | 0 (0%) | -.054 | 19/91 (20.9%) | 2239 | 31 (1.38%) |

*Note.* Summary of item-level absolute fit (RMSD > .10), local item dependence ($|Q3| > .20$), and person-fit outliers ($|Zh| > 3$). Person-fit statistics ($Zh$) calculated per Drasgow et al. (1985) via *mirt*'s personfit() function; values of $|Zh| > 3$ denote statistically atypical response vectors.
*Abbreviations:* GRM = Graded Response Model; RMSD = Root Mean Square Deviation.

**Supplemental Table S4**

*Flagged CHRT-PR$_{16}$ Item Pairs with Absolute Q3 Greater Than .20 Top 10 of 25 by Magnitude*

| **Item 1** | **Item 2** | $Q_3$ |
|---|---|---|
| Irritable/easily angered (12) | Overreacts with anger/rage (13) | .660 |
| Acts without thinking (10) | Decisions on impulse (11) | .614 |
| Thoughts how to kill self (15) | Has a plan to kill self (16) | .526 |
| Can do nothing right (3) | Everything turns out wrong (4) | .491 |
| Thoughts of killing self (14) | Thoughts how to kill self (15) | .342 |
| Decisions on impulse (11) | Overreacts with anger/rage (13) | .319 |
| Decisions on impulse (11) | Irritable/easily angered (12) | .306 |
| Acts without thinking (10) | Irritable/easily angered (12) | .256 |
| Acts without thinking (10) | Overreacts with anger/rage (13) | .252 |
| Thoughts of killing self (14) | Has a plan to kill self (16) | .244 |

*Note.* Item pairs exhibiting residual local dependence evaluated via Yen's *Q*3 statistic following unidimensional GRM estimation. Values of $|Q3| > .20$ indicate residual correlation exceeding expected unidimensional model assumptions.

*Abbreviations:* GRM = Graded Response Model; CHRT-PR$_{16}$ = Concise Health Risk Tracking Parent Report.

**Supplemental Table S5**

*CHRT-PR$_{14}$ GRM item parameters (excludes items 12 & 13)*

| Item | a | b1 | b2 | b3 | b4 | Discrimination |
|---|---|---|---|---|---|---|
| Never going to get better (1) | 2.71 | 0.13 | 1.02 | 1.73 | 2.60 | very high |
| No future (2) | 3.69 | 0.57 | 1.36 | 1.92 | 2.57 | very high |
| Can do nothing right (3) | 2.71 | 0.20 | 1.10 | 1.74 | 2.56 | very high |
| Everything turns out wrong (4) | 2.93 | 0.23 | 1.17 | 1.72 | 2.74 | very high |
| No one to depend on (5) | 3.82 | 0.64 | 1.45 | 1.96 | 2.78 | very high |
| People cared for are gone (6) | 2.55 | 0.69 | 1.54 | 2.10 | 2.69 | very high |
| Wishes suffering were over (7) | 3.19 | 0.75 | 1.33 | 1.86 | 2.55 | very high |
| No reason to live (8) | 6.22 | 0.94 | 1.53 | 1.98 | 2.65 | very high |
| Wishes to sleep, not wake (9) | 4.96 | 0.98 | 1.59 | 1.96 | 2.49 | very high |
| Acts without thinking (10) | 1.24 | −0.49 | 0.42 | 1.25 | 3.19 | moderate |
| Decisions on impulse (11) | 1.07 | −0.84 | 0.26 | 1.24 | 3.36 | moderate |
| Thoughts of killing self (14) | 5.56 | 1.02 | 1.55 | 1.88 | 2.56 | very high |
| Thoughts how to kill self (15) | 5.30 | 1.05 | 1.57 | 2.00 | 2.42 | very high |
| Has a plan to kill self (16) | 4.75 | 1.18 | 1.72 | 2.27 | 2.72 | very high |

*Note.* Item parameters estimated using a unidimensional Graded Response Model (GRM) on the complete-case baseline sample ($N = 2{,}239$), excluding irritability items 12 and 13. Parameter $a$ represents item discrimination (slopes), categorized according to Baker and Kim's (2017) criteria. Parameters $b1$ through $b4$ represent item difficulty thresholds along the latent trait θ scale.
*Abbreviations:* GRM = Graded Response Model; CHRT-PR$_{14}$ = Concise Health Risk Tracking 14-Item Parent Report.

**Supplemental Table S6**

*Item Fit RMSD for CHRT-PR$_{14}$ (Excluding Items 12 and 13 from CHRT-PR$_{16}$)*

| Item | RMSD |
|---|---|
| Decisions on impulse (11) | .086 |
| Acts without thinking (10) | .082 |
| Can do nothing right (3) | .029 |
| Everything turns out wrong (4) | .029 |
| Never going to get better (1) | .026 |
| Wishes suffering were over (7) | .020 |
| No future (2) | .015 |
| No one to depend on (5) | .013 |
| People cared for are gone (6) | .013 |
| Thoughts how to kill self (15) | .012 |
| Wishes to sleep, not wake (9) | .008 |
| Thoughts of killing self (14) | .008 |
| No reason to live (8) | .006 |
| Has a plan to kill self (16) | .006 |

*Note.* Item-level Root Mean Square Deviation (RMSD) absolute model fit statistics computed manually per Yamamoto et al. (2013) across decile bins of EAP θ trait estimates. Values of RMSD ≤ .10 indicate acceptable item-level model fit. Zero items across either scale length exceeded the .10 misfit boundary. *Abbreviations:* EAP = Expected A Posteriori; GRM = Graded Response Model; RMSD = Root Mean Square Deviation.

**Supplemental Table S7**

*Flagged CHRT-PR$_{14}$ Item Pairs with Absolute Q3 Greater Than .20 Top 5 of 19*

| Item 1 | Item 2 | $Q_3$ |
|---|---|---|
| Acts without thinking (10) | Decisions on impulse (11) | .635 |
| Can do nothing right (3) | Everything turns out wrong (4) | .501 |
| Thoughts how to kill self (15) | Has a plan to kill self (16) | .476 |
| Thoughts of killing self (14) | Thoughts how to kill self (15) | .292 |
| Never going to get better (1) | No future (2) | .200 |

*Note.* Item pairs exhibiting residual local dependence evaluated via Yen's *Q*3 statistic following unidimensional GRM estimation. Values of $|Q3| > .20$ indicate residual correlation exceeding expected unidimensional model assumptions.
*Abbreviations:* GRM = Graded Response Model; CHRT-PR$_{14}$ = Concise Health Risk Tracking Parent Report 14-item.

**Supplemental Table S8**

*Cross-Validated Scoring-Method Comparison (Unit-Weighted vs. EAP Theta) on the 2-Week Retest Holdout (N = 170)*

| Criterion Measure | $r_{UW}$ | $r_{EAP}$ | Diff | Cohen's *q* | Zou 95% CI | *p* (Holm) |
|---|---|---|---|---|---|---|
| ***CHRT-PR$_{16}$*** | | | | | | |
| PSC-17 Total | .684 | .559 | .124 | .205 | [.073, .189] | < .001 |
| PSC-17 Attention | .485 | .367 | .118 | .145 | [.057, .186] | .004 |
| PSC-17 Externalizing | .488 | .371 | .116 | .144 | [.055, .184] | .004 |
| PSC-17 Internalizing | .687 | .630 | .057 | .101 | [.007, .115] | .452 |
| Aggression/Disruptive Dx | .253 | .176 | .077 | .081 | [.010, .144] | .452 |
| NQLS | -.582 | -.523 | -.059 | .085 | [-.121, -.002] | .618 |
| Attended Therapy | .169 | .107 | .062 | .063 | [-.006, .130] | 1.00 |
| Rx Medication | .079 | .037 | .042 | .042 | [-.026, .110] | 1.00 |
| Depression Dx | .133 | .136 | -.004 | .003 | [-.071, .064] | 1.00 |
| VQIDS | .445 | .425 | .021 | .025 | [-.042, .084] | 1.00 |
| OTC/Herbal/Supplement | .144 | .134 | .009 | .010 | [-.059, .077] | 1.00 |
| N-Gauge | .418 | .391 | .027 | .032 | [-.037, .091] | 1.00 |
| ***CHRT-PR$_{14}$*** | | | | | | |
| PSC-17 Total | .633 | .513 | .120 | .180 | [.064, .187] | < .001 |
| PSC-17 Attention | .453 | .325 | .128 | .151 | [.064, .198] | .002 |
| PSC-17 Externalizing | .414 | .315 | .098 | .114 | [.033, .167] | .065 |
| PSC-17 Internalizing | .676 | .619 | .057 | .098 | [.004, .116] | .571 |
| Aggression/Disruptive Dx | .216 | .155 | .061 | .063 | [-.009, .130] | 1.00 |
| NQLS | -.568 | -.520 | -.049 | .068 | [-.112, .010] | 1.00 |
| Attended Therapy | .124 | .066 | .058 | .059 | [-.012, .128] | 1.00 |
| Rx Medication | .068 | .029 | .039 | .039 | [-.032, .109] | 1.00 |
| Depression Dx | .093 | .118 | -.025 | .025 | [-.095, .045] | 1.00 |
| VQIDS | .418 | .412 | .007 | .007 | [-.059, .073] | 1.00 |
| OTC/Herbal/Supplement | .107 | .118 | -.011 | .011 | [-.081, .059] | 1.00 |
| N-Gauge | .361 | .374 | -.013 | .015 | [-.080, .053] | 1.00 |

*Note.* Both scores were computed on the strict-matched Time-2 retest holdout (N = 170) using GRM item parameters estimated on the full Time-1 sample. Holm correction was applied across all 24 comparisons

jointly. Positive Diff and Cohen's q values favor the unit-weighted score throughout (NQLS is reverse-scored, so its negative Diff also favors the unit-weighted score in absolute magnitude).
*Abbreviations:* Dx = diagnosis; EAP = Expected A Posteriori (Item Response Theory theta score); GRM = Graded Response Model; N-Gauge = Parent-Rated Global Impairment Scale; NQLS = Nationwide Quality of Life Scale (reverse-scored); OTC = over-the-counter/herbal/supplement; PSC-17 = Pediatric Symptom Checklist-17; rEAP = Pearson correlation coefficient using EAP theta score; rUW = Pearson correlation coefficient using unit-weighted raw sum score; Rx = prescription; VQIDS = Very Quick Inventory of Depressive Symptomatology (5-item parent proxy).

**Supplemental Table S9**

*Comparison of Candidate GAMLSS Distributional Models for CHRT-PR$_{16}$ and CHRT-PR$_{14}$ Continuous Norming*

| Candidate Model | Distribution Type | df | BIC | $Z_3$ (Skewness) | $Z_4$ (Kurtosis) | Agostino $K^2$ |
|---|---|---|---|---|---|---|
| ***CHRT-PR$_{16}$ Parent Report (16 Items; N = 2,235)*** | | | | | | |
| **ZINBI** | Zero-Inflated Negative Binomial (Type I) | 7.24 | 14,992.40 | 10.05 (p = .61) | 36.88 (p < .001) | 46.94 (p < .001) |
| **ZANBI** | Zero-Adjusted Negative Binomial (Hurdle) | 7.40 | 15,004.79 | 9.61 (p = .65) | 36.41 (p < .001) | 46.02 (p = .01) |
| **NBI** | Negative Binomial (Type II) | 6.22 | 15,103.01 | 51.87 (p < .001) | 41.56 (p < .001) | 93.43 (p < .001) |
| ***CHRT-PR$_{14}$ Parent Report (14 Items; N = 2,235)*** | | | | | | |
| **ZINBI** | Zero-Inflated Negative Binomial (Type I) | 7.13 | 13,938.94 | 8.20 (p = .77) | 28.67 (p = .01) | 36.88 (p = .06) |
| **ZANBI** | Zero-Adjusted Negative Binomial (Hurdle) | 7.27 | 13,953.40 | 8.11 (p = .78) | 28.12 (p = .01) | 36.23 (p = .07) |
| **NBI** | Negative Binomial (Type II) | 6.16 | 14,011.11 | 29.06 (p = .01) | 45.40 (p < .001) | 74.46 (p < .001) |

*Note.* Candidate distributional regression models were fitted using Generalized Additive Models for Location, Scale and Shape (GAMLSS) regressing total scores on penalized B-spline terms for age, sex main effects, and sex-by-age interactions. All models used a log link for location (mu) and dispersion (sigma) submodels, with constant zero-inflation (nu) where applicable. Model selection prioritized the Bayesian Information Criterion (BIC), where lower values indicate superior fit penalizing for model complexity. Local residual diagnostics were evaluated across age bands using worm plots and Q-statistics: $Z_3$ assesses residual skewness, $Z_4$ assesses residual kurtosis, and Agostino $K^2$ provides a combined omnibus test of residual normality. NBI failed severely across both scale lengths due to unmodeled zero-inflation driving significant residual skewness ($p < .01$). ZINBI and ZANBI performed similarly on local residual diagnostics, but ZINBI was selected as the final norming model for both scales due to superior global deviance and BIC calibration.

*Abbreviations:* Agostino $K^2$ = D'Agostino omnibus test for residual normality; BIC = Bayesian Information Criterion; CHRT-PR$_{14}$ = Concise Health Risk Tracking 14-Item Parent Report; CHRT-PR$_{16}$ = Concise Health Risk Tracking 16-Item Parent Report; GAMLSS = Generalized Additive Models for Location, Scale and Shape; NBI = Negative Binomial Type II distribution; ZANBI = Zero-Adjusted Negative Binomial distribution; ZINBI = Zero-Inflated Negative Binomial distribution; $Z_3$ = Q-statistic for residual skewness; $Z_4$ = Q-statistic for residual kurtosis.

**Supplemental Figure S1**

*De-Trended Quantile Residual Worm Plots Across Developmental Age Subgroups for the CHRT-$PR_{16}$ GAMLSS ZINBI Norming Model*

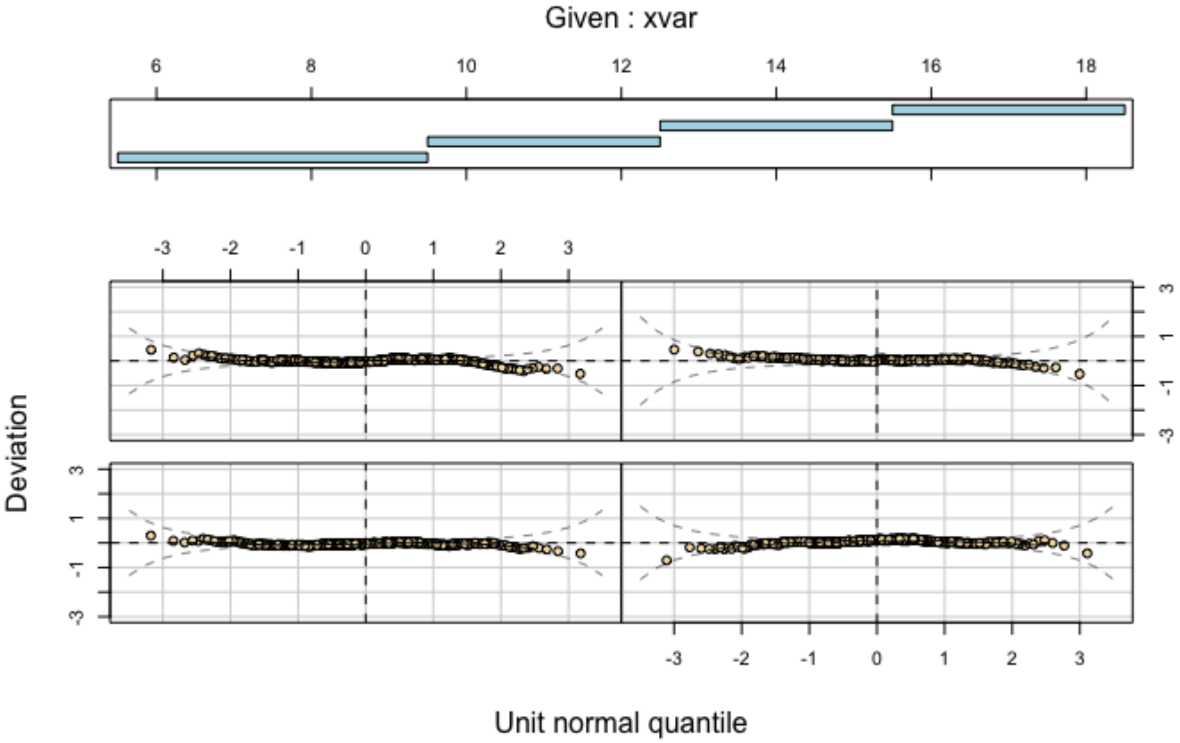


*Note.* Trellis de-trended Q-Q residual plots (worm plots) produced for the final Zero-Inflated Negative Binomial (ZINBI) Generalized Additive Models for Location, Scale and Shape (GAMLSS) continuous norming model for the CHRT-$PR_{16}$. Data are conditioned across four developmental age intervals: 6.0 to 9.5 years ($n$ = 673), 9.5 to 12.5 years ($n$ = 534), 12.5 to 15.5 years ($n$ = 661), and 15.5 to 18.0 years ($n$ = 367). The horizontal axis plots unit normal quantiles and the vertical axis plots residual deviation from normality. Dashed parabolic curves represent point-wise 95% confidence bounds. Residual points scattering randomly around the horizontal zero line within the 95% confidence intervals demonstrate adequate local residual calibration across developmental stages.
*Abbreviations:* CHRT-$PR_{16}$ = Concise Health Risk Tracking 16-Item Parent Report; GAMLSS = Generalized Additive Models for Location, Scale and Shape; ZINBI = Zero-Inflated Negative Binomial distribution.

**Supplemental Figure S2**

*De-Trended Quantile Residual Worm Plots Across Developmental Age Subgroups for the CHRT-PR$_{14}$ GAMLSS ZINBI Norming Model*

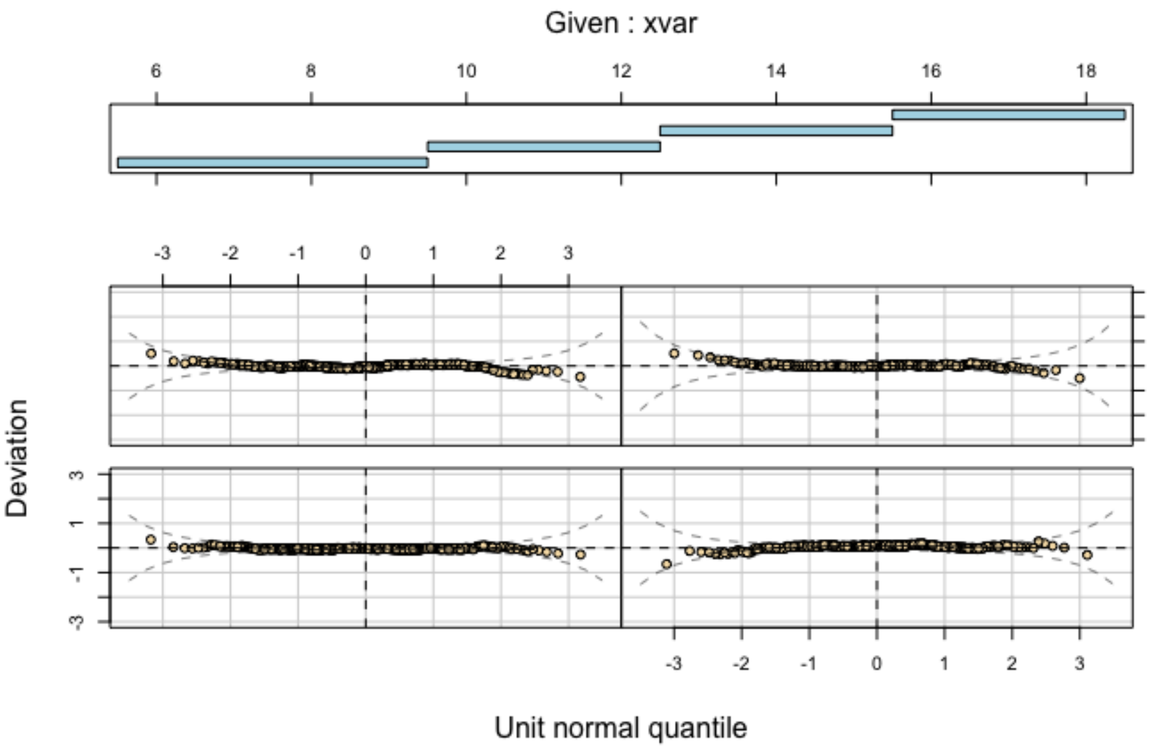


*Note.* Trellis de-trended Q-Q residual plots (worm plots) produced for the final Zero-Inflated Negative Binomial (ZINBI) Generalized Additive Models for Location, Scale and Shape (GAMLSS) continuous norming model for the CHRT-PR$_{14}$. Data are conditioned across four developmental age intervals: 6.0 to 9.5 years ($n$ = 673), 9.5 to 12.5 years ($n$ = 534), 12.5 to 15.5 years ($n$ = 661), and 15.5 to 18.0 years ($n$ = 367). The horizontal axis plots unit normal quantiles and the vertical axis plots residual deviation from normality. Dashed parabolic curves represent point-wise 95% confidence bounds. Residual points scattering randomly around the horizontal zero line within the 95% confidence intervals demonstrate adequate local residual calibration across developmental stages.
*Abbreviations:* CHRT-PR$_{14}$ = Concise Health Risk Tracking 14-Item Parent Report; GAMLSS = Generalized Additive Models for Location, Scale and Shape; ZINBI = Zero-Inflated Negative Binomial distribution.

**Supplemental Figure S3**

*Bland-Altman Plots of Test-retest Stability with Johnson-Neyman Regions of Significance for CHRT-$PR_{16}$*

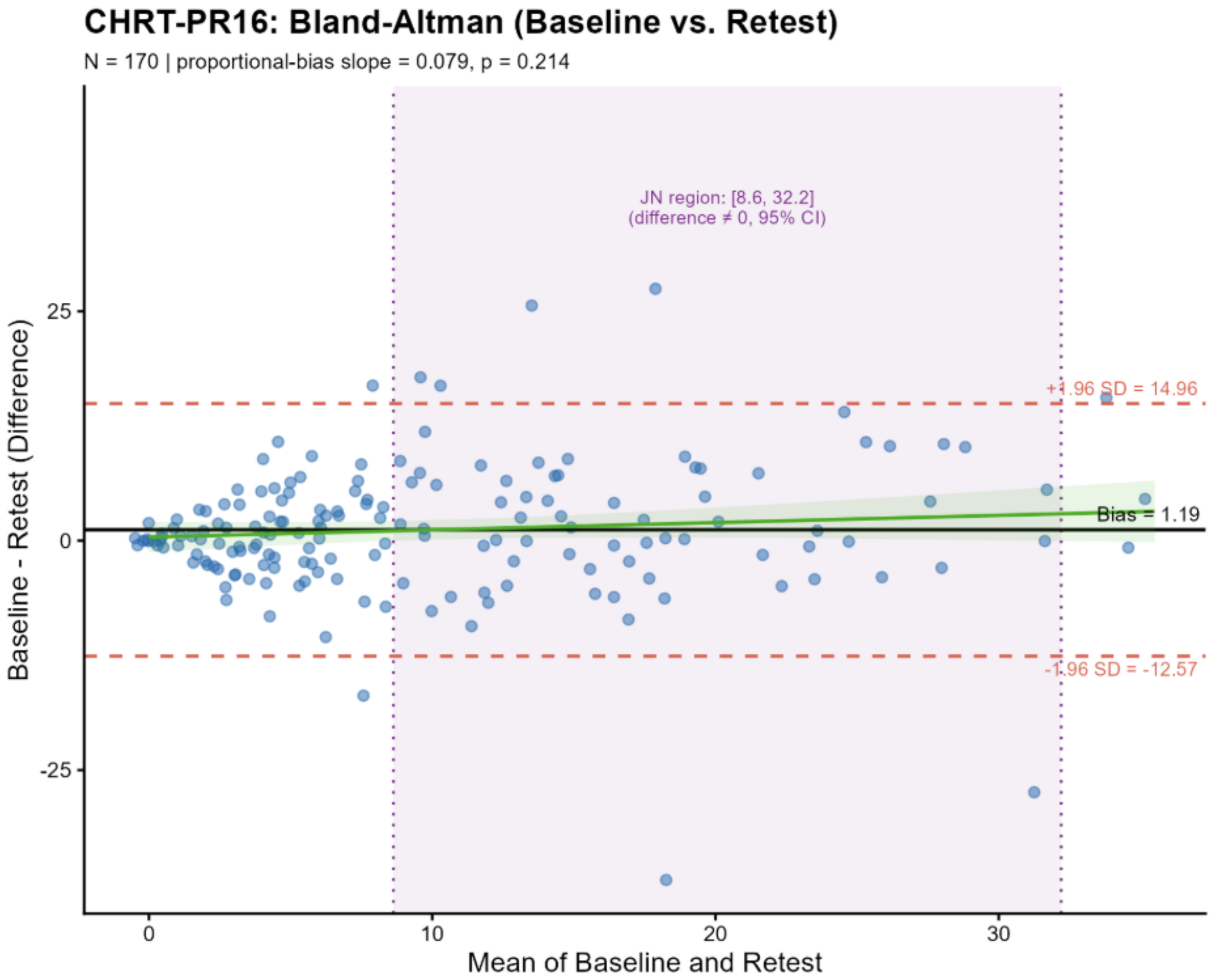


*Note.* Bland–Altman agreement plot and Johnson–Neyman region of significance evaluating 2-week test-retest score stability and proportional bias for the CHRT-PR16 in the retest holdout sample ($n$ = 170). The horizontal axis displays the mean of baseline (Time 1) and retest (Time 2) total scores; the vertical axis displays individual score differences (Time 1 − Time 2). The solid horizontal line represents mean baseline-to-retest score change (1.19 points, $dz$ = 0.17), and dashed horizontal lines denote 95% limits of agreement (±1.96 $SD$). The fitted linear regression line demonstrates no statistically significant proportional bias across severity levels ($b$ = 0.08, $p$ = .21). Shaded vertical region highlights the Johnson–Neyman region of non-significance across observed scores [8.6, 32.2], confirming score stability across non-extreme clinical severity ranges.
*Abbreviations:* CHRT-$PR_{16}$ = Concise Health Risk Tracking 16-Item Parent Report.

**Supplemental Figure S4**

*Bland-Altman Plots of Test-retest Stability with Johnson-Neyman Regions of Significance for CHRT-PR$_{14}$*

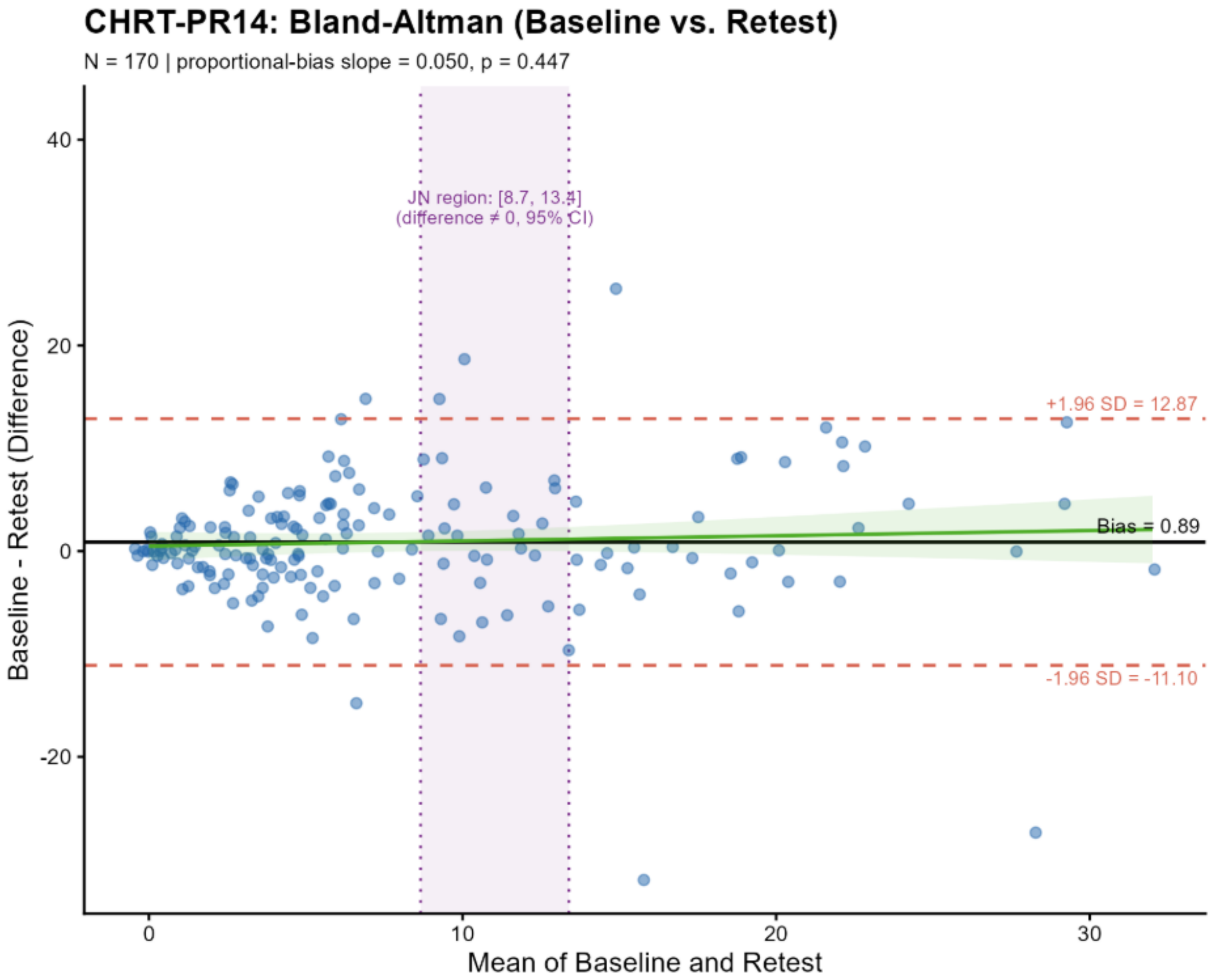


*Note.* Bland–Altman agreement plot and Johnson–Neyman region of significance evaluating 2-week test-retest score stability and proportional bias for the CHRT-PR14 in the retest holdout sample ($n$ = 170). The horizontal axis displays the mean of baseline (Time 1) and retest (Time 2) total scores; the vertical axis displays individual score differences (Time 1 − Time 2). The solid horizontal line represents mean baseline-to-retest score change (0.89 points, $dz$ = 0.15), and dashed horizontal lines denote 95% limits of agreement (±1.96 $SD$). The fitted linear regression line demonstrates no statistically significant proportional bias across severity levels ($b$ = 0.05, $p$ = .45). Shaded vertical region highlights the Johnson–Neyman region of non-significance across observed scores [8.7, 13.4].

*Abbreviations:* CHRT-PR$_{14}$ = Concise Health Risk Tracking 14-Item Parent Report.
